# A toolkit for quantification of biological age from blood-chemistry and organ-function-test data: BioAge

**DOI:** 10.1101/2021.08.28.21262759

**Authors:** Dayoon Kwon, Daniel W. Belsky

**Affiliations:** Department of Epidemiology, Fielding School of Public Health, University of California, Los Angeles, Los Angeles, CA, 90095, USA; Robert N. Butler Columbia Aging Center, Mailman School of Public Health, Columbia University, New York, NY, 10032, USA; Department of Epidemiology, Mailman School of Public Health, Columbia University, New York, NY, 10032, USA

**Keywords:** Biological age, Geroscience, CALERIE, Biomarkers, Aging, Healthspan

## Abstract

Methods to quantify biological aging are emerging as new measurement tools for epidemiology and population science and have been proposed as surrogate measures for healthy lifespan extension in geroscience clinical trials. Publicly available software packages to compute biological aging measurements from DNA methylation data have accelerated dissemination of these measures and generated rapid gains in knowledge about how different measures perform in a range of datasets. Biological age measures derived from blood chemistry data were introduced at the same time as the DNA methylation measures and, in multiple studies, demonstrate superior performance to these measures in prediction of healthy lifespan. However, their dissemination has been slow by comparison, resulting in a significant gap in knowledge. We developed a software package to help address this knowledge gap. The BioAge R package, available for download at GitHub (http://github.com/dayoonkwon/BioAge), implements three published methods to quantify biological aging based on analysis of chronological age and mortality risk: Klemera-Doubal Biological Age, PhenoAge, and homeostatic dysregulation. The package allows users to parametrize measurement algorithms using custom sets of biomarkers, to compare the resulting measurements to published versions of the Klemera-Doubal method and PhenoAge algorithms, and to score the measurements in new datasets. We applied BioAge to safety lab data from the CALERIE™ randomized controlled trial, the first-ever human trial of long-term calorie restriction in healthy, non-obese adults, to test effects of intervention on biological aging. Results contribute evidence that CALERIE intervention slowed biological aging. BioAge is a toolkit to facilitate measurement of biological age for geroscience.

## Introduction

Biological aging is the gradual and progressive decline in system integrity that occurs with advancing chronological age [1]. Processes of biological aging begin with accumulation of cellular-level changes that increase vulnerability of tissues and organs to loss of function, ultimately causing disease, disability, and death [2, 3]. Experiments with animals show these processes can be modified, extending healthy lifespan for worms, flies, and mice [4, 5]. The emerging field of geroscience is focused on translating these therapies to extend healthy lifespan in humans [6, 7]. Key to these translational efforts is the development of biomarkers that can detect effects of treatments that slow or reverse biological aging.

Development of biomarkers of aging has a long history and remains a work in progress [8]. For geroscience, aging biomarkers are needed because it will take decades to establish whether treatments extend healthy lifespan in humans [9, 10]. Biomarker measurements, by contrast, have potential provide early tests of treatment effectiveness over timescales of months or years.

There is still no gold standard biomarker of aging. Among those showing most promise are a family of algorithms applied to DNA methylation data that estimate a person’s biological age or mortality risk [11-18]. These biomarkers first emerged early in the last decade and since then have undergone rapid refinement, increasing in their reliability and predictive power [19]. Many clinical and cohort studies are now conducting DNA methylation analysis of stored biospecimens. Using publicly available tools, e.g. https://horvath.genetics.ucla.edu/html/dnamage/, research teams around the world are using these datasets to compute DNA methylation measures of aging and advance the science.

A second set of promising aging biomarkers are algorithms derived from blood chemistries and other clinical data. Although these algorithms based on clinical parameters are as or more predictive of disease, disability, and mortality as compared to DNA methylation measures [20-22] and show evidence of sensitivity to a range of causes hypothesized to accelerate aging [23-26], they have received much less research attention. One barrier to wider integration of these clinical-data algorithms into aging research is a lack of software for computing these measures in new datasets. A further barrier is that many studies will include several but not all of the clinical markers included in a particular algorithm. Unlike DNA methylation datasets, which are generated from a single multiplex array and include the same measurements across studies, datasets of clinical markers are assembled from multiple assays. As a result, a study may be missing one or another of the markers included in an algorithm.

To address these barriers, we present a novel R package, “BioAge”, which is currently programmed to implement three methods to quantify biological aging: Klemera-Doubal method (KDM) Biological Age [27], PhenoAge [13], and homeostatic dysregulation [28]. The package has two sets of functions. One set of functions allows the user to develop new versions of the KDM Biological Age, PhenoAge and homeostatic dysregulation measures using biomarker sets of their own choosing and data from the US Health and Nutrition Examination Surveys (NHANES). This set of functions (1) trains new algorithms using data from NHANES III; and (2) compares the new algorithms to published versions using data from NHANES IV. The second set of functions allows the user to train new versions of the algorithms and then apply them to test data of the users choosing. Together, these functions enable users to develop new versions of published algorithms that are customized to the biomarkers available within their own datasets. BioAge was designed as an easy-to-use package that only requires a set of available clinical markers as inputs, enabling researchers to compute the biological aging measures and evaluate the performance of the biological age algorithms. Here, we provide an example of BioAge implementation using data from a randomized controlled trial, CALERIE. The CALERIE trial tested the effects of two-years of caloric restriction in a sample of healthy, non-obese adults. We use the BioAge package to compute measures of biological aging at pre-intervention baseline and at 12- and 24-month follow-up assessments. We then use the computed measures to evaluate the effect of CALERIE intervention on biological aging.

## Methods

The BioAge package develops algorithms to measure biological aging from a user-specified list of biomarkers based on three published methods: the Klemera-Doubal Method Biological Age (KDM BA) [27], the PhenoAge [13], and homeostatic dysregulation (HD) [28]. The package includes datasets for training and testing algorithms from the US Health and Nutrition Examination Surveys (https://wwwn.cdc.gov/nchs/nhanes/Default.aspx). The package is available on GitHub (http://github.com/dayoonkwon/BioAge) and is licensed under the GNU General Public License v3.0.

The package contains two sets of functions. The first set of functions (i) apply published methods to generate biological age algorithms based on a user-specified list of biomarkers using the National Health and Nutrition Examination Surveys (NHANES) III dataset as a training sample; and (ii) compares these new algorithms to one another and to published algorithms using NHANES IV dataset as a test sample. These functions are labeled with the suffix “_nhanes” (kdm_nhanes, phenoage_nhanes, hd_nhanes). The second set of functions apply published methods to generate biological age algorithms based on a user-specified list of biomarkers using a user-specified training dataset and projects these new algorithms onto a user-specified test dataset. These functions make it possible to train new algorithms using the NHANES datasets and then to project these algorithms onto new test datasets. These functions are labeled with the suffix “_calc” (kdm_calc, phenoage_calc, hd_calc).

The following sections introduce the three methods of calculating biological age, the NHANES data, and the comparative analysis performed in the _nhanes functions.

### Biological aging measures

The BioAge package calculates biological aging measures using three methods: KDM BA, PhenoAge, and HD [13, 27, 29]. These biological aging measures are patient-level measures that combine information from multiple clinical biomarkers to quantify aging-related deficits in system integrity [27, 29, 30]. We selected these three methods based on previous literature and published evidence for links with morbidity, mortality, and indicators of healthspan in young and older populations [13, 27, 30-34].

#### KDM Biological Age

An individual’s KDM BA prediction corresponds to the chronological age at which her/his physiology would be approximately normal. KDM BA older than chronological age indicates an advanced state of biological aging and increased risk for disease, disability, and mortality. KDM BA younger than biological age indicates delayed biological aging and reduced risk for disease, disability, and mortality.

The KDM BA algorithm is derived from a series of regressions of individual biomarkers on chronological age in a reference population. The equation takes information from *n* number of regression lines of chronological age regressed on *n* biomarkers:

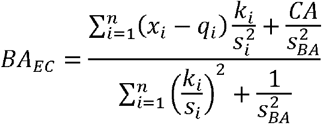

where *x* is the value of biomarker *i* measured for an individual. For each biomarker *i*, the parameters *k, q*, and *s* are estimated from a regression of chronological age on the biomarker in the reference sample. *k, q*, and *s* are the regression intercept, slope, and root mean squared error, respectively. *s*_*BA*_ is a scaling factor equal to the square root of the variance in chronological age explained by the biomarker set in the reference sample. CA is chronological age. In the kdm_nhanes function in BioAge package, the reference sample is NHANES III nonpregnant participants aged 30-75 years. Algorithm parameters are estimated separately for men and women.

#### PhenoAge

An individual’s PhenoAge prediction corresponds to the chronological age at which their mortality risk would be approximately normal in a reference population. PhenoAge older than chronological age indicates an advanced state of biological aging and increased risk for disease, disability, and mortality. PhenoAge younger than biological age indicates delayed biological aging and reduced risk for disease, disability, and mortality.

The PhenoAge algorithm is derived from multivariate analysis of mortality hazards [34, 35]. The original PhenoAge algorithm was constructed from elastic-net Gompertz regression of mortality on 42 biomarkers in the NHANES III [13]. This analysis selected nine biomarkers and chronological age as a parsimonious model. This model is used to compute a mortality prediction score. The mortality prediction score is then converted into a biological age value by matching the elastic-net model predicted score with mortality scores from a univariate Gompertz regression including only chronological age as a predictor. The chronological age at which the univariate model prediction matches the elastic-net model prediction is assigned as the biological age.

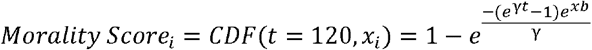

where *xb* represents the linear combination of biomarkers from the fitted model. γ is an ancillary parameter to be estimated from the data and *t* denotes time (here in units of months). Thus, *CDF(t = 120, x*_*i*_) denotes the probability that the *i*^*th*^ individual will die within the next 120 months. The mortality score is then converted to a PhenoAge value. This conversion is made based on a univariate Gompertz regression of the mortality hazard including only chronological age:

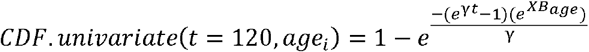

estimates the probability that the *i*^*th*^ individual will die within the next 120 months as follows *CDF*.*univariate(120, age*_*i*_) where *age*_*i*_ is the chronological age of the *i*^*th*^ individual. In the “phenoage_nhanes” function, a set of biomarkers specified by the users is used to form the mortality prediction score in place of the original elastic-net model.

#### Homeostatic Dysregulation

An individual’s HD value corresponds to how different their physiology is from a healthy reference. Higher values of HD indicate an advanced state of biological aging and increased risk for disease, disability, and mortality. Lower values of homeostatic dysregulation indicate delayed biological aging and reduced risk for disease, disability, and mortality.

HD is computed as the Mahalanobis distance [36] for a set of biomarkers relative to a reference sample. The Mahalnobis distance equation [36] takes the form:

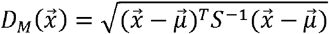

where *x* is a multivariate observation (all the biomarker values for an individual) and μ is the equivalent-length vector of reference sample means for each variable. *S* is the reference sample variance-covariance matrix for the variables. If all variables are uncorrelated then this is equivalent to scaling each biomarker by its variance and then summing the squared deviance for an observation:

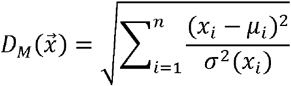

where *n* is the number of biomarkers and σ^*2*^*(x*_*i*_) is the variance in the *i*^*th*^ biomarker. In the hd_nhanes function, we specify the reference sample to be NHANES III nonpregnant participants aged 20-30 years for whom all user-selected biomarkers fall within the clinically normal range. For analysis, all biomarkers are standardized to have mean=0, SD=1 separately for men and women based on this reference sample. This approach computes homeostatic dysregulation relative to a young, healthy sample, following the approach we have used previously [24, 33, 37].

## NHANES

NHANES is an ongoing nationally representative, cross-sectional survey conducted by the US Centers for Disease Control and Prevention. NHANES administers questionnaires during in-home interviews and conducts health examinations, including blood draws, in a mobile examination center. Details of recruitment procedures and study design are available from the Center for Disease Control and Prevention (https://www.cdc.gov/nchs/nhanes/index.htm). We compiled demographic, socioeconomic, functional performance, biomarker, and mortality data from adults aged 20-90 years participating in the NHANES III (1988-1994) and IV (1999-2018).

For analysis, we excluded biomarker outliers by computing sex-specific mean of standard deviations and dropping values more than five standard deviations from the sex-specific mean. Biomarkers with skewed distributions were log-transformed. Details on biomarker measurements are available from the NHANES website (https://www.cdc.gov/nchs/nhanes/index.htm).

For several biomarkers, measurement methods changed during the 1999-2016 interval when NHANES IV data used by the package were collected. The package uses data normalized to account for these changes in methodology: Creatinine values from the 1999-2000 and 2005-2006 NHANES were corrected according to the analytical notes posted by NHANES (https://www.cdc.gov/nchs/nhanes/index.htm). High sensitivity C-reactive protein assays from the 2015-2016 NHANES were posted in units mg/L and were divided by 10 to match units in previous waves. Bone alkaline phosphatase values from the 1999-2000 NHANES were adjusted according to published equations. Measurement methods for plasma fasting glucose changed in the 2005-2006, 2007-2008, and 2015-216 NHANES. Values were adjusted to be comparable across years using multiple regression equations (https://www.cdc.gov/nchs/nhanes/index.htm). Measurement methods for insulin changed in the 1999-2000, 2003-2004, 2005-2006, 2011-2012, and 2013-2014 NHANES. Values were adjusted to be comparable across years using multiple regression equations (https://www.cdc.gov/nchs/nhanes/index.htm).

In the BioAge package, data from NHANES III and NHANES IV are loaded as the datasets NHANES3 and NHANES4. The NHANES4 dataset also contains computed values of KDM Biological Age and PhenoAge based on the original versions of those algorithms published by Levine and colleagues [13, 30].

### Comparison of Biological Aging Measures

The package’s _nhanes functions conduct a series of analyses to compare KDM BA, PhenoAge, and HD algorithms generated with user-specified sets of biomarkers with one another and with versions of KDM BA and PhenoAge algorithms published previously. All algorithms are trained in NHANES III data. Comparative analysis is conducted using NHANES IV data. Thus, training and test samples are distinct from one another. Analyses proceed in four steps. First, biological aging measures are correlated with chronological age. Second, biological aging measures are correlated with one another. Third, biological aging measures are tested for association with healthspan-related characteristics: mortality, disability physical function, and self-rated health. Finally, a set of analyses tests socioeconomic patterning of biological aging algorithms.

Below, the measures included in the validation analysis are described briefly, followed by details of the analysis. Complete details on all measurements are available from the NHANES website (https://www.cdc.gov/nchs/nhanes/index.htm).

#### Mortality

NHANES III and NHANES 1999-2014 are independent cohorts with different lengths of follow-up for mortality. Participants’ survival status and cause of death were determined through probabilistic matching to the death certificates form the National Death Index recorded through 2015 [38]. For analysis, we used information on aging-related mortality from diseases of the heart, malignant neoplasms, chronic lower respiratory disease, cerebrovascular disease, Alzheimer’s disease, Diabetes mellitus, nephritis, nephrotic syndrome, and nephrosis.

#### Disability, Physical Functioning and Self-rated Health

We analyzed associations of biological aging measures with counts of limitations to activities of daily living (ADLs), walk speed, grip strength, and self-rated health. ADLs were measured as a count of functional impairments across 19 activities. Walk speed was measured in NHANES 1999-2002 from participants aged 50 and older. Measures were taken as time in seconds to walk 20 feet. Values were log-transformed for analysis. Grip strength was measured in NHANES 2011-2014 using a hand dynamometer. Values were averaged across three trials. We analyzed values for the dominant hand. To account for differences in distributions between men and women, values were transformed to have M=0, SD=1 within sex. Self-rated general health was assessed from a survey item with five response categories ranging from excellent to poor.

#### Socioeconomic Circumstances

Socioeconomic circumstances measures included education, annual family income, and poverty income ratio. Education was categorized into five categories: less than 9^th^ grade, 9-11^th^ grade, high school graduate/GED, some college, college graduate or higher. Annual family income was self-reported as one of 11 income categories ranging from less than $5,000 to over $75,000. Poverty income ratio was calculated based on family income and the poverty threshold based on family size.

### Analysis of Validation Measures

For analysis, KDM BA and PhenoAge values were differenced from chronological age values and then standardized to have M=0, SD=1 separately for men and women. HD values were log transformed and then standardized to have M=0, SD=1 separately for men and women. The package tests associations of biological aging measures with mortality (survival time) using Cox proportional hazards models to estimate Hazard Ratios (HR). The package tests associations of biological aging measures with counts of ADLs, log-walk-test time, grip strength, and self-rated health using linear regression models. For linear regression analysis, outcome variables are standardized to have M=0, SD=1. Coefficients reported are interpretable on a Pearson’s r scale. The package tests associations of socioeconomic circumstances measures with biological aging measures using linear regression. In these models, measures of socioeconomic circumstances are specified as independent variables and biological aging measures are specified as dependent variables. Socioeconomic circumstance measures are standardized to M=0, SD=1 for analysis so that effect-sizes are denominated in terms of a 1 SD unit improvement in socioeconomic circumstances. Analysis includes NHANES IV participants for whom biological aging measures can be calculated and for whom any validation data are available. Models are reported for the full analysis sample with covariate adjustment for chronological age and sex and for samples stratified by sex, race, and chronological age (under/over 65y).

### Analysis

We compared original KDM BA and PhenoAge algorithms with algorithms composed with the new biomarker set in the NHANES IV dataset. First, two sets of “plot_” functions create comparative scatter plots using Pearson correlations. “plot_ba” function tests associations of chronological age with biological aging measures. “plot_baa” function tests associations among biological aging measures. In this function, KDM BA and PhenoAge were computed as the difference between biological age and chronological age. These biological age advancement (BAA) values were then standardized to have mean = 0, SD = 1 separately for men and women within the analysis sample.

Three sets of “table_” functions create regression tables with full sample and stratifying by gender, race, and age groups. “table_surv” function tests associations of biological aging measures with mortality. This function uses Cox proportional hazard models to estimate hazard ratios. “table_health” function tests associations of biological aging measures with functional performance measures. Biological aging measures were independent variables. Functional performance measures were dependent variables and standardized to have mean = 0, SD = 1 for analysis. “table_ses” function tests association of socioeconomic circumstance measures with biological aging measures. Socioeconomic circumstance measures were independent variables and standardized to have mean = 0, SD = 1 for analysis. Biological aging measures were dependent variables. In table_health and table_ses functions, linear regression is used to compute standardized beta coefficients (interpretable as Pearson’s r).

### Results

#### Part 1. Parameterization of KDM, PhenoAge, and HD biological aging measures for the CALERIE Trial dataset using NHANES III

We previously analyzed KDM and HD biological aging measures in CALERIE using a biomarker set based on the original KDM algorithm published by Levine [33, 39]. We used the _nhanes functions of the BioAge package to train new KDM, PhenoAge, and HD algorithms in the NHANES III data and then used the _calc functions to project the algorithms onto the CALERIE data. We conducted three sets of analyses: (i) using the same biomarker set included in our original CALERIE analysis (hereafter, “CALERIE Original”); (ii) using a biomarker set based on the original PhenoAge algorithm published by Levine (Levine et al., 2018) (hereafter “V1”); and (iii) using a biomarker set composed of those included in the CALERIE Original and V1 sets (hereafter “V2”). Biomarkers included in the three sets of measures and their correlations with chronological age in the NHANES IV test data and the CALERIE sample at baseline are reported in **Table S2.1 Panel A**.

#### Part 2: Validation of new KDM, PhenoAge, and HD algorithms in NHANES IV data

The several biological aging measures were correlated with one another in the NHANES IV test data (**Figure 1**). To analyze correlations among measures, we first differenced KDM and PhenoAge measures from chronological age to calculate age-advancement values. Correlations among the biological aging measures in NHANES IV ranged from 0.39-0.96 (**Fig. 1, Fig. S1.2**).

**Fig. 1.**
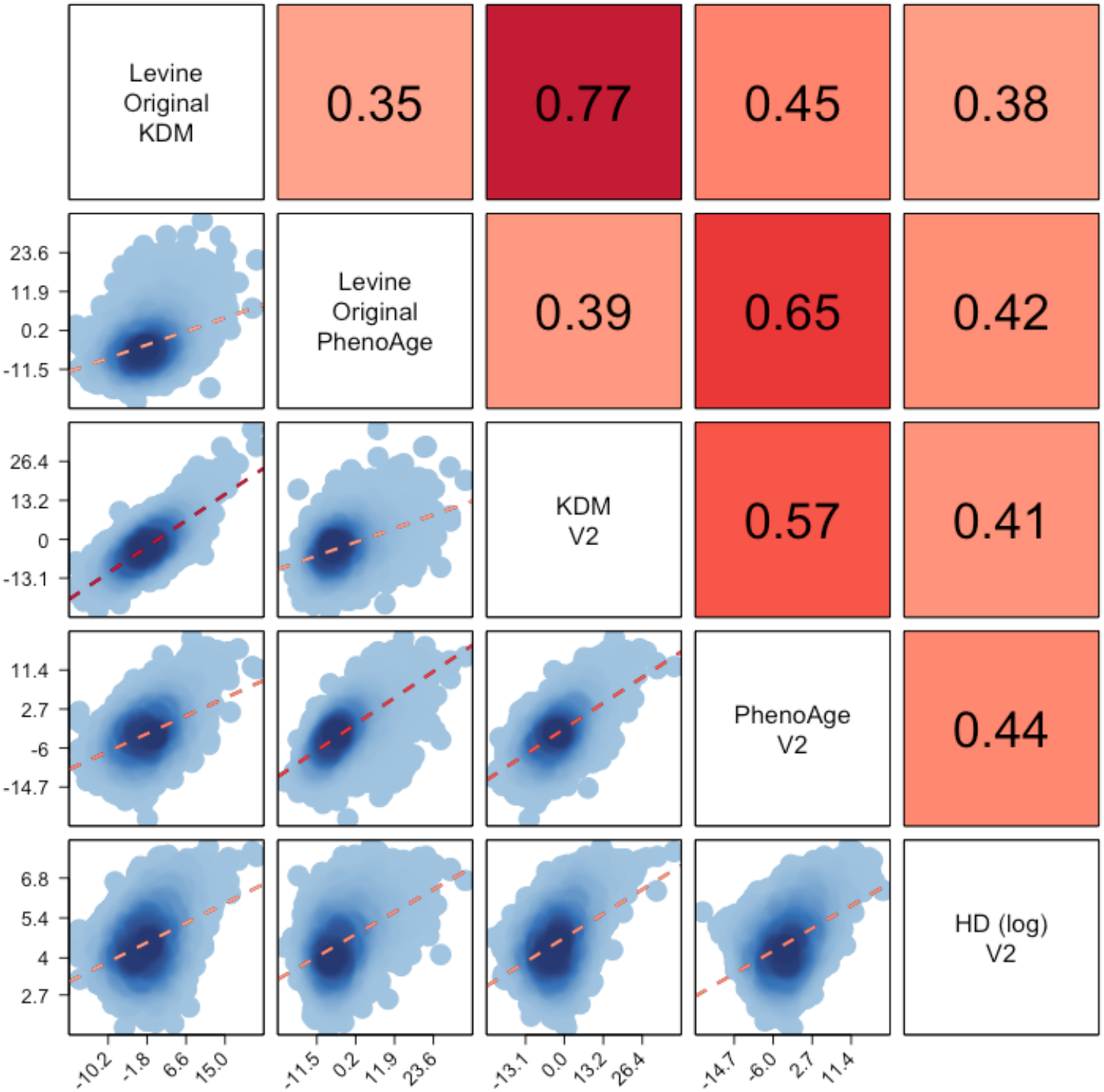
Correlations of published versions of Klemera-Doubal method (KDM) Biological Age and PhenoAge with versions of KDM Biological Age, PhenoAge, and a measure computed using the homeostatic dysregulation method based on a modified set of biomarkers. The figure plots data from NHANES IV generated with the _nhanes commands within the BioAge package. All measures were developed from analysis of NHANES III and computed using data from NHANES IV. KDM Biological Age and PhenoAge values were differenced from chronological age for analysis. The “Levine Original” KDM algorithm was composed from chronological age and 10 biomarkers: albumin, alkaline phosphatase, blood urea nitrogen, creatinine, C-reactive protein, cytomegalovirus optical density, HbA1C, total cholesterol, systolic blood pressure, and forced expiratory volume in 1 second (FEV_1_). The “Levine Original” PhenoAge was composed from chronological age and 9 biomarkers: albumin, alkaline phosphatase, creatinine, C-reactive protein, fasting glucose, white blood cell count, lymphocyte percentage, mean cell volume, and red cell distribution width. The “V2” versions of the KDM, PhenoAge, and HD algorithms included chronological age and 12 biomarkers: albumin, alkaline phosphatase, blood urea nitrogen, creatinine, C-reactive protein, HbA1C, total cholesterol, uric acid, white blood cell count, lymphocyte percentage, mean cell volume, and red cell distribution width

The several biological aging measures were associated with mortality risk in the NHANES IV test sample. Compared to the KDM and PhenoAge algorithms first proposed by Levine, the CALERIE-Original and V1 algorithms were somewhat less predictive of mortality and more-weakly correlated with self-rated health and measures of physical functioning; the V2 algorithms, which included biomarkers from both the CALERIE-Original and V1 sets, performed better, although still not as well as the original PhenoAge. Complete results from analysis of the NHANES IV data produced by the plot_ and table_ functions of the BioAge package are compiled in **SI Section I**. These include analysis of association with chronological age, intercorrelation with other measures of aging, including the original KDM and PhenoAge algorithms proposed by Levine [13, 39], and additional association analysis of self-rated health, and physical functioning, and socioeconomic factors. The V2 algorithm consistently performed better in these criterion validity analyses relative to the CALERIE-Original and V1 algorithms. Therefore, we retained the V2 algorithm for CALERIE analysis. (For completeness of documentation, results for CALERIE analysis of the CALERIE-Original and V1 algorithms are reported in **SI Section II**.)

#### Part 3. CALERIE Analysis

The CALERIE Trial randomized n=220 non-obese midlife adults to two years of 25% caloric restriction (CR) or ad libitum (AL) diet, the control condition. Most participants in the CR intervention group did not achieve the prescribed dose of caloric restriction; the average percent CR over the two years was about half the prescribed dose [40]. Nevertheless, participants in the CR group lost significant weight over the first 12 months of the trial and maintained this weight loss over the second 12-month interval. They also experienced a range of physiological changes indicating improved cardiometabolic health [41]. We previously reported that the CR group demonstrated slower biological aging as compared to the AL group based on versions of the KDM BA and HD algorithms trained in data from NHANES 2007-11 [33]. PhenoAge has not yet been analyzed in CALERIE. Characteristics of the CALERIE participants included in analysis and values of the biological aging measures in the CR-intervention and AL-control groups are reported in **Table 1**.

**Table1.**
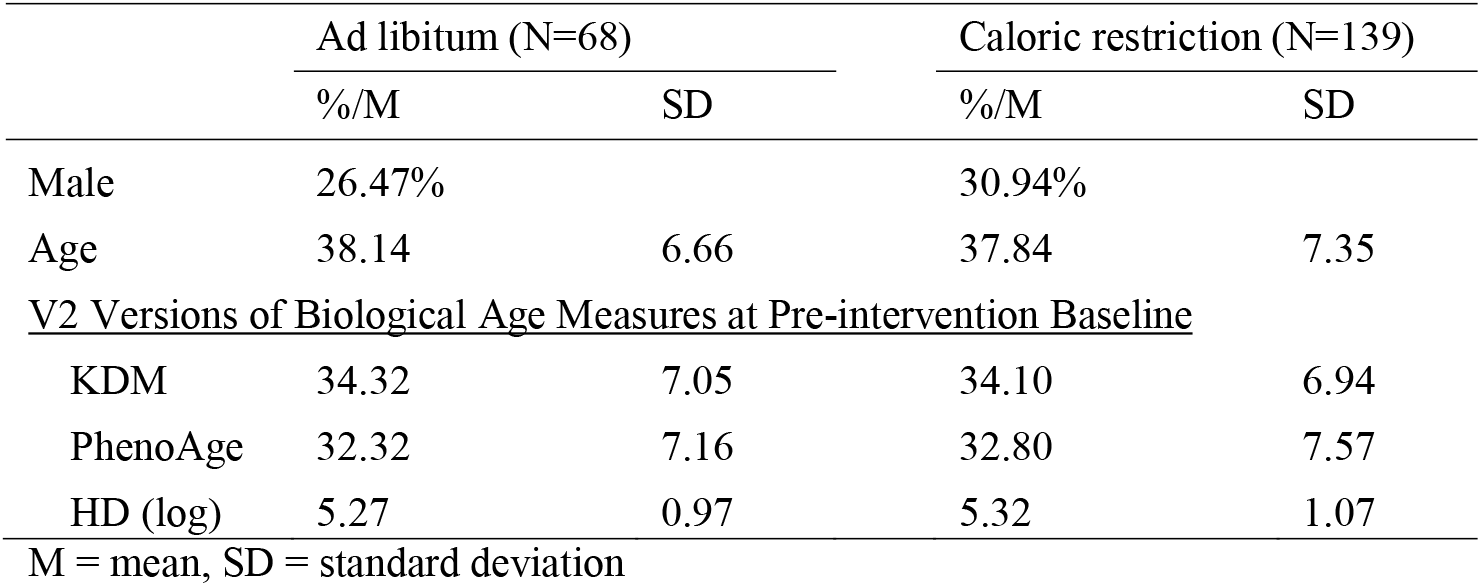
Characteristics of CALERIE Trial Participants included in analysis

|  | Ad libitum (N=68) |  | Caloric restriction (N=139) |  |
| --- | --- | --- | --- | --- |
|  | %/M | SD | %/M | SD |
| Male | 26.47% |  | 30.94% |  |
| Age | 38.14 | 6.66 | 37.84 | 7.35 |
| <u>V2 Versions of Biological Age Measures at Pre-intervention Baseline</u> |  |  |  |  |
| KDM | 34.32 | 7.05 | 34.10 | 6.94 |
| PhenoAge | 32.32 | 7.16 | 32.80 | 7.57 |
| HD (log) | 5.27 | 0.97 | 5.32 | 1.07 |
| M = mean, SD = standard deviation |  |  |  |  |

CALERIE participants’ chronological age and V2 versions of the KDM BA and PhenoAge measures were correlated (Pearson r range =0.81-0.90, **Fig. 2**). CALERIE participants’ V2 HD values were not correlated with their chronological ages. At baseline, there was little difference in biological aging measures between CR and AL groups. Participants’ biological aging measures were slightly younger than their chronological ages (chronological age mean =39; KDM BA mean=35; PhenoAge mean=34). This difference may reflect the sampling frames used for CALERIE and the NHANES, volunteer bias, and that CALERIE participants were selected to be in good health, whereas the NHANES sample represented the general US population. CALERIE sample baseline summary statistics for biological aging measures are reported in **Table S2.2**. Intercorrelations of biological aging measures are graphed in **Fig. S2.2**.

**Fig. 2.**
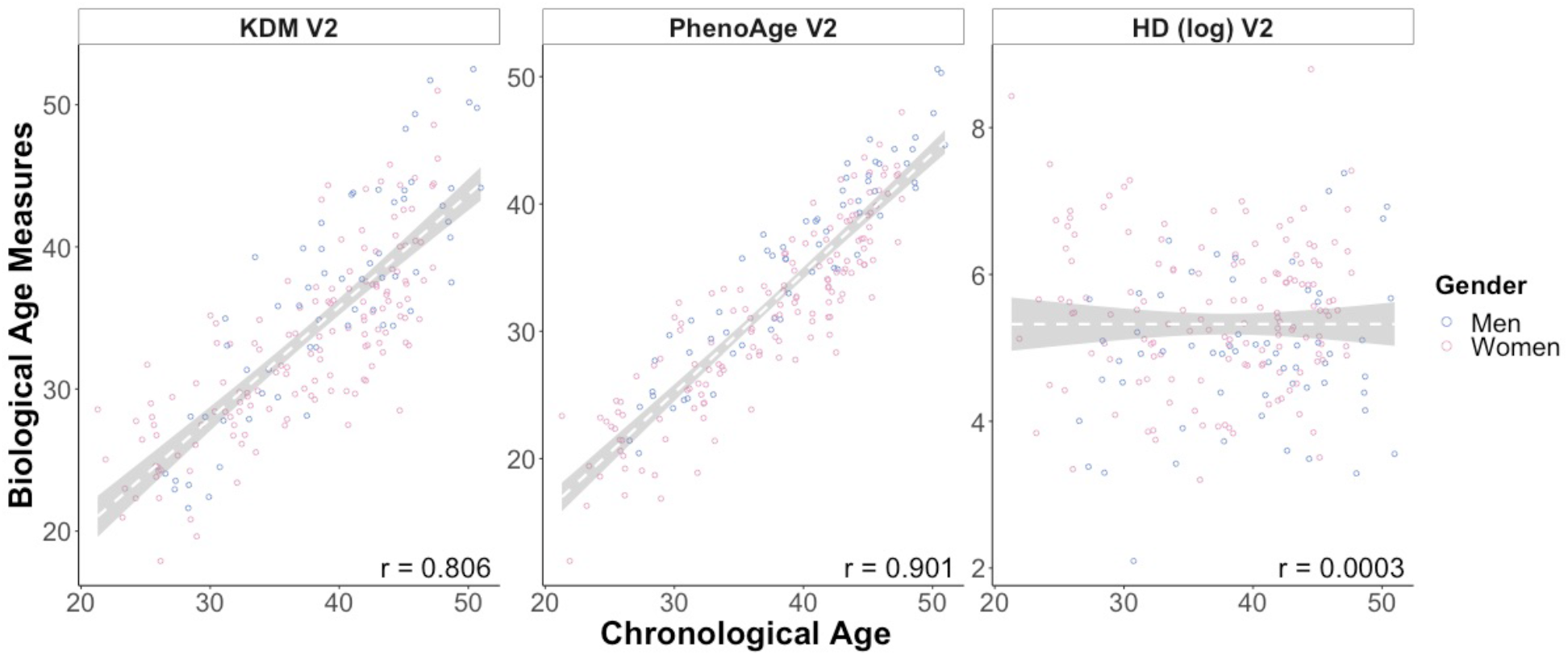
Associations of Klemera-Doubal method (KDM) Biological Age, PhenoAge, and homeostatic dysregulation (HD) measures of biological age with chronological age among participants in the CALERIE Trial at pre-intervention baseline. The figure plots pre-intervention baseline values of the three biological aging measures against chronological age for men (blue) and women (pink) participating in the CALERIE Trial (n=207)

We tested the hypothesis that CALERIE interventions slowed biological aging using mixed-effects growth models, including participant-level random intercepts and slopes. The model took the form

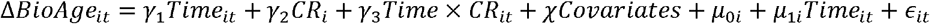

Where ΔBioAge_it_ is change in the BA measure from baseline for individual “i” at time “t”, *γ*_1_ estimates annual change in BA for ad libitum-arm participants, *γ*_2_ estimates any baseline difference BA between participants in AL and CR arms of the trial, *γ*_3_ estimates the difference in annual change in BA between AL-arm and CR-arm participants, *χ* is a vector of covariates, and *µ*_0*i*_ and *µ*_1*i*_ are the random intercepts and slopes estimated for each individual “i”. The coefficient *γ*_3_ tests the hypothesis that biological aging was slowed for participants randomized to the CR arm of the trial. All models included sex and baseline age as covariates. The mixed-effects regression analysis included 611 observations of 220 individuals.

Across follow-up, CALERIE participants randomized to the trial’s CR arm experienced slower or reversed biological aging as compared to AL arm participants as measured by V2 KDM BA and PhenoAge. The average change from baseline in biological aging measures is plotted for AL and CR participants in **Fig. 3**. CALERIE participants randomized to the AL control group experienced an increase in V2 KDM BA of 0.69 “years” per 12-month follow-up (95% CI [0.21, 1.16]). In contrast, for participants randomized to CR, KDM BA decreased (b=-0.2 95% CI [-0.55, 0.14] “years” per 12-month follow-up interval). For PhenoAge, both AL and CR groups experienced an increase over time, but the increase was slower for the CR group (AL b=0.83 [0.46-1.21], CR b=0.17 [-0.11, 0.44]; p-value for test of difference =0.006). HD was unchanged across follow-up both AL- and CR-group participants. Regression model results are reported in **Table 2**.

**Fig. 3.**
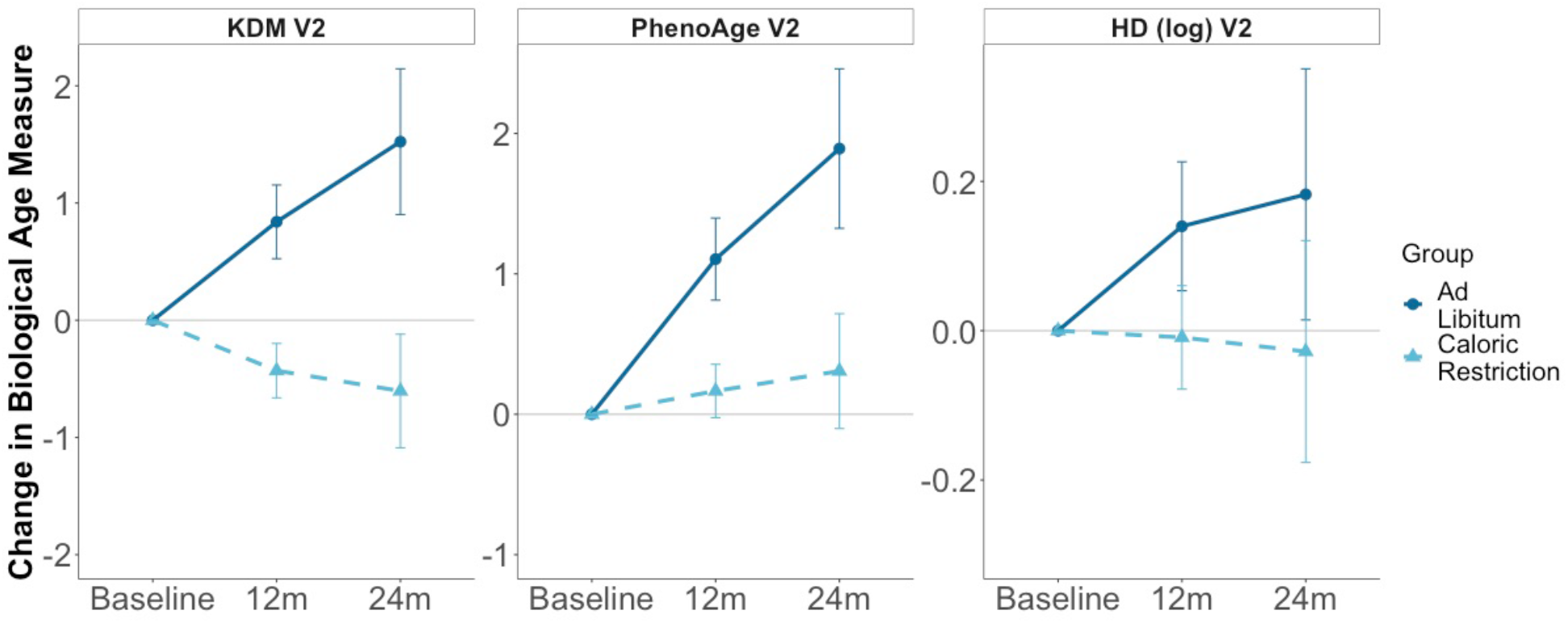
Change in Klemera-Doubal method (KDM) Biological Age, PhenoAge, and homeostatic dysregulation (HD) from Baseline to 12- and 24-month follow-ups in the ad libitum (dark blue dots) and caloric-restriction (light blue triangles) groups of the CALERIE trial. The figure plots predicted values and 95% confidence intervals estimated from mixed-effects growth models for participants in the ad libitum control group (dark blue circles, solid line) and caloric restriction intervention group (light blue triangles, dashed line). Values of KDM Biological Age and PhenoAge are denominated in years. Values of HD are denominated in log units

**Table 2.**
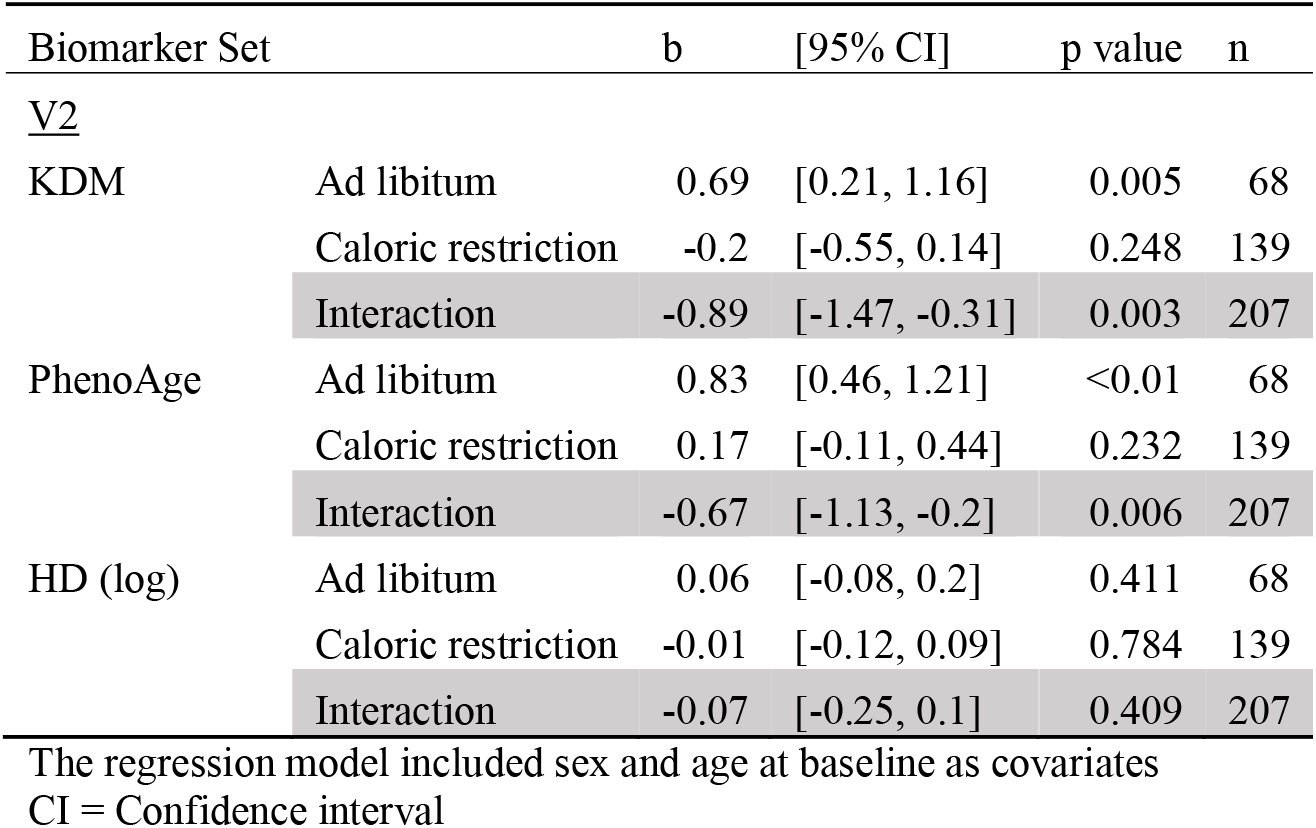
Estimated annual change in KDM, PhenoAge, and HD biological aging measures from baseline through 24-month follow-up in Ad libitum- and Caloric Restriction groups in the CALERIE Randomized Trial

## Discussion

Quantification of biological aging is emerging as a novel approach to investigating how a range of exposures and interventions may influence risk for chronic disease, disability, and mortality [8]. Because the aging process is ongoing from at least reproductive maturity and may begin even earlier [42], biological aging measures have potential to detect signs of risk decades before disease processes are established [31]. Measurements of biological aging based on algorithm-defined composites of clinical parameters are relatively understudied in this growing research area. These measures are equally or more predictive of morbidity and mortality as compared to better-studied measures based on molecular data, including telomere length and DNA methylation clocks [20-22, 24]; they are variable in apparently healthy young adults [23, 31]; and they are sensitive to risk exposures that shorten healthy lifespan and to interventions that slow aging in animals [23, 24, 33, 43, 44]. Importantly, the data needed to calculate these algorithm-based measures are routinely collected during clinical care and health research: routine blood chemistries, complete blood counts, and assessments of lung function and blood pressure. They therefore represent an un-tapped reservoir of information about aging processes within many existing datasets. We developed an R package to aid investigators in integrating these measurements into existing datasets that addresses key challenges that may have slowed their adoption.

The BioAge R package is an easy to install tool that can implement the Klemera-Doubal [27], PhenoAge [35], and homeostatic dysregulation [28] methods following the approach we have used in previous work [24, 33, 43, 45]. The package has three strengths. First, it eases implementation of published biological age algorithms in biomarker datasets. Second, it allows for parameterization of new algorithms using published methods based in existing or new datasets. Third, when new algorithms are composed of biomarkers included in the NHANES database, it enables head-to-head comparison with the original published versions of the biological age algorithms to evaluate comparative criterion and construct validity.

We demonstrated the BioAge package by applying it to calculate biological age values from laboratory data collected in the CALERIE randomized controlled trial, the first human randomized controlled trial of long-term calorie restriction [40]. We previously used data from NHANES 2007-2010 to develop versions of the KDM and HD algorithms to test effects of CALERIE intervention on biological aging [33]. Building on that analysis, we used the BioAge package to compute new versions of the PhenoAge, KDM, and HD algorithms based on (a) the original set of markers used in our earlier paper; (b) a set of markers matched as closely as possible to the Levine PhenoAge algorithm; and (c) a combined set of measures included in the first two sets. We used NHANES III data to train these algorithms. In comparative validation analysis using data from NHANES IV (1999-2018), the algorithm with the combined set of biomarkers performed the best in analysis predicting morbidity and mortality. We applied this algorithm to the CALERIE data to test intervention effects on biological aging. Consistent with our earlier analysis, we found that CALERIE intervention slowed biological aging as measured by the KDM and PhenoAge algorithms. However, in contrast to our previous analysis, we detected no effect on biological aging as measured by the HD algorithm. This difference in result likely reflects differences in the training samples (NHANES 2007-2010 in our original analysis and NHANES III, conducted during 1989-1994, in this analysis). Future applications of the HD algorithm may be better served by matching years of the training sample to the years of collection of the test data.

## Conclusions

Measurements of biological aging derived from clinical parameters, including routine blood chemistries, complete blood count data, and blood pressure and lung function testing, represent a powerful approach to investigating etiology of individual differences in aging and evaluating interventions to increase healthspan. The BioAge R package facilitates implementation of training and testing of three common, validated methods to compute biological age measurements from these types of data.

## Data Availability

All data used in this manuscript are publicly available. Data from the US National Health and Nutrition Examinations Surveys are available from the US Centers for Disease Control and Prevention at https://wwwn.cdc.gov/nchs/nhanes/Default.aspx. Data from the CALERIE Trial are available from the CALERIE Research Network at https://calerie.duke.edu/samples-data-access-and-analysis.

https://calerie.duke.edu/samples-data-access-and-analysis

## Declarations

### Code availability

All the code used in these analyses is covered by GPL-3.0 License and is available from GitHub: https://github.com/dayoonkwon/BioAge.

### Authors’ contributions

DWB and DK conceived the research and designed the software and analysis. DK wrote the software, conducted the analysis, and produced the figures. DWB and DK wrote the manuscript. Both authors had access to the data.

### Ethics approvals

Not applicable.

### Consent to participant

Not applicable.

### Consent for publication

Not applicable.

