## Supplemental Information for "A toolkit for quantification of biological age from blood-chemistry and organ-function-test data: BioAge"

**Supplementary Information**

*I. Analysis of the NHANES IV data using the BioAge package*

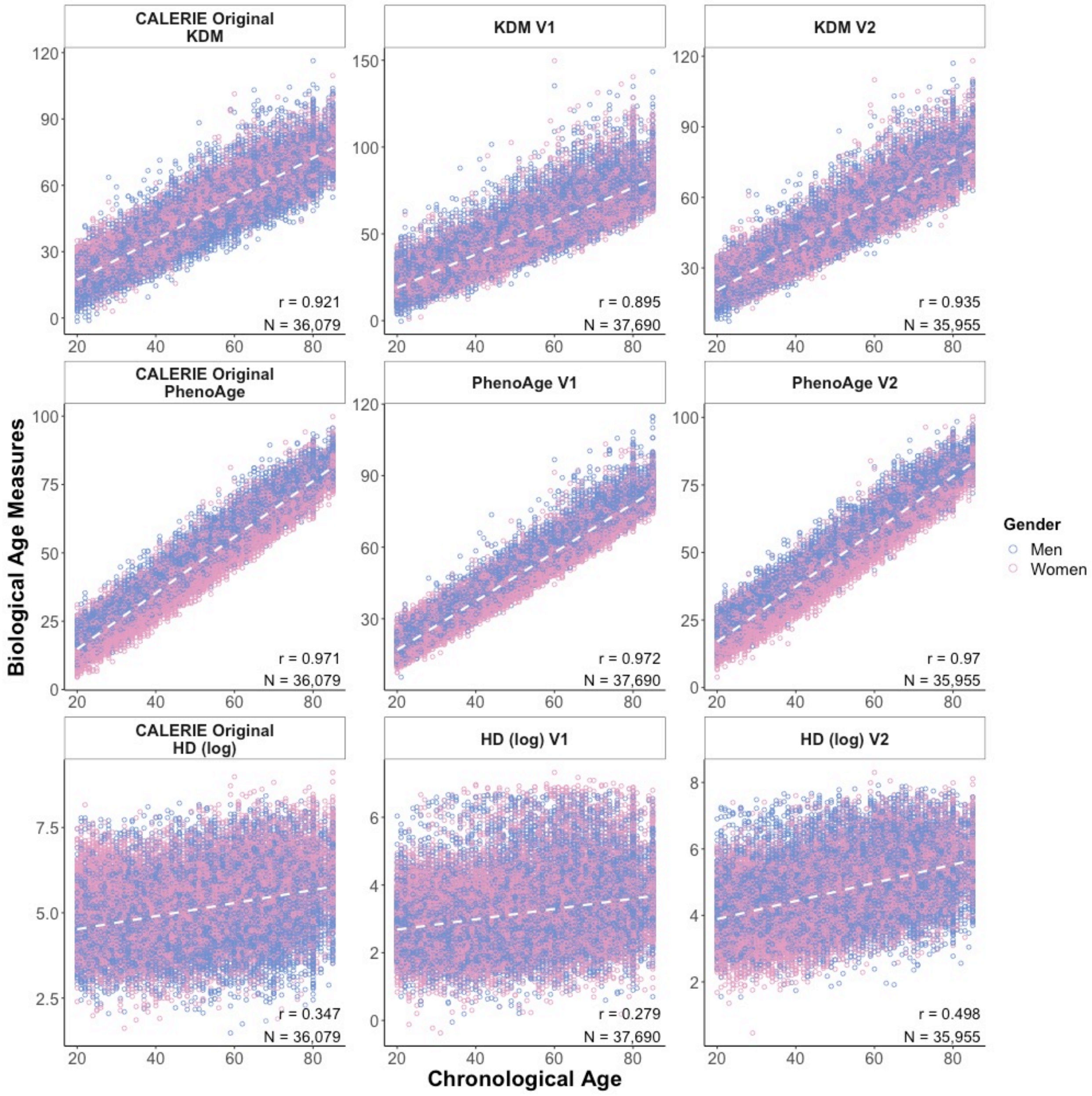


**Fig. S1.1** KDM, PhenoAge, and HD biological aging measures plotted against chronological age for participants in the NHANES IV

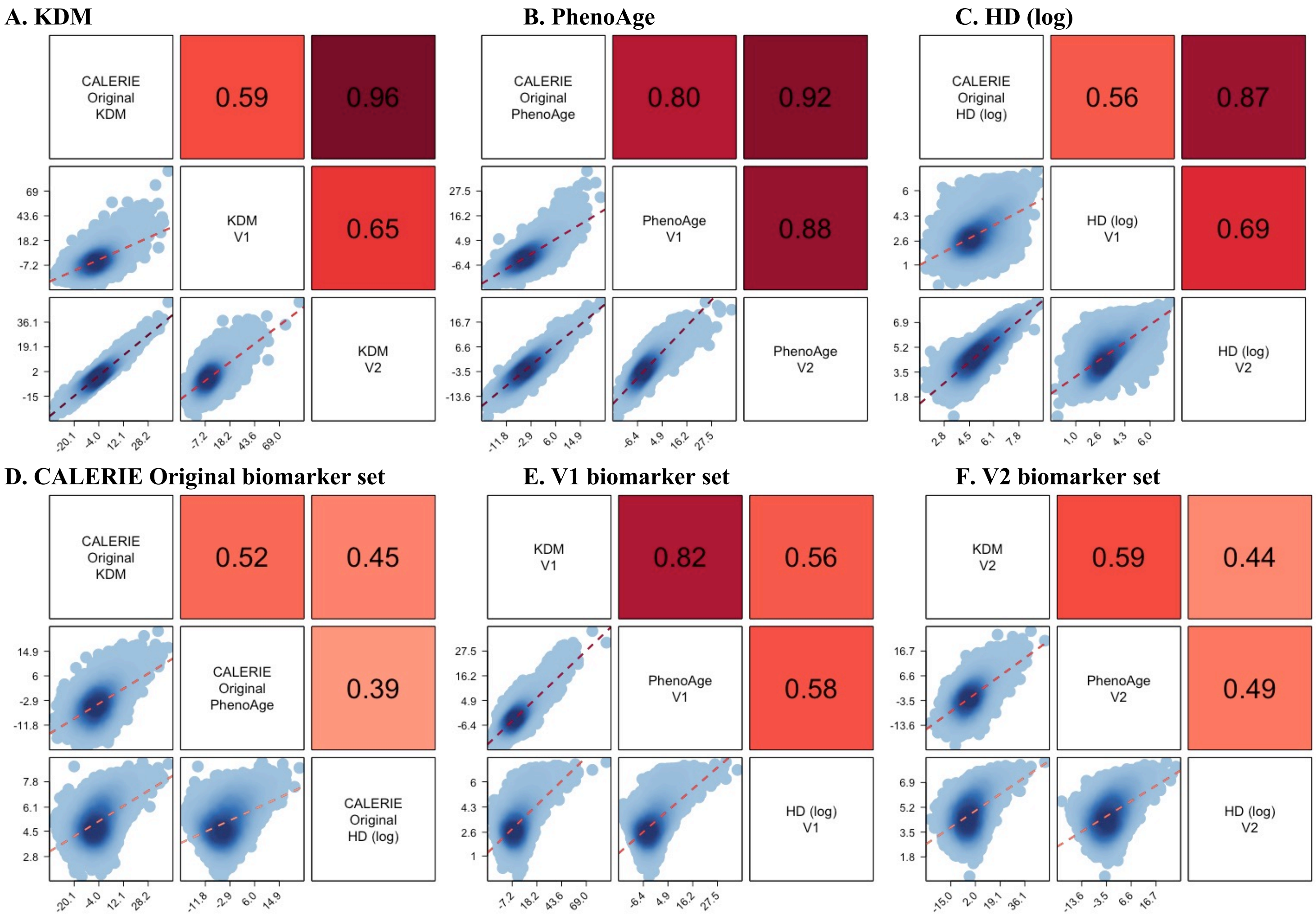


**Fig. S1.2** A matrix of association plots among KDM, PhenoAge, and HD biological aging measures in the NHANES IV

**Table S1.1** Associations of biological aging measures with mortality in the NHANES IV

| | Original | | CALERIE Original | | | V1 | | | V2 | | |
|---|---|---|---|---|---|---|---|---|---|---|---|
| | KDM | PhenoAge | KDM | PhenoAge | HD (log) | KDM | PhenoAge | HD (log) | KDM | PhenoAge | HD (log) |
| | Hazard Ratio (95% CI) | | | | | | | | | | |
| Full Sample | | | | | | | | | | | |
| n | 8,234 | 27,837 | 26,688 | 26,688 | 26,688 | 27,904 | 27,904 | 27,904 | 26,580 | 26,580 | 26,580 |
| BA | 1.36 | 1.47 | 1.24 | 1.39 | 1.33 | 1.35 | 1.39 | 1.39 | 1.26 | 1.43 | 1.44 |
| | (1.2, 1.55) | (1.42, 1.51) | (1.2, 1.29) | (1.34, 1.45) | (1.28, 1.39) | (1.3, 1.39) | (1.34, 1.43) | (1.34, 1.44) | (1.22, 1.31) | (1.37, 1.49) | (1.38, 1.51) |
| Stratified by Gender | | | | | | | | | | | |
| Men | | | | | | | | | | | |
| n | 4,114 | 13,421 | 12,907 | 12,907 | 12,907 | 13,453 | 13,453 | 13,453 | 12,879 | 12,879 | 12,879 |
| BA | 1.44 | 1.44 | 1.26 | 1.38 | 1.3 | 1.37 | 1.35 | 1.37 | 1.28 | 1.41 | 1.4 |
| | (1.23, 1.69) | (1.38, 1.5) | (1.2, 1.32) | (1.31, 1.45) | (1.24, 1.37) | (1.31, 1.43) | (1.29, 1.41) | (1.31, 1.44) | (1.22, 1.34) | (1.34, 1.49) | (1.32, 1.49) |
| Women | | | | | | | | | | | |
| n | 4,120 | 14,416 | 13,781 | 13,781 | 13,781 | 14,451 | 14,451 | 14,451 | 13,701 | 13,701 | 13,701 |
| BA | 1.23 | 1.52 | 1.22 | 1.41 | 1.38 | 1.32 | 1.46 | 1.41 | 1.24 | 1.46 | 1.53 |
| | (0.98, 1.54) | (1.45, 1.6) | (1.15, 1.3) | (1.32, 1.5) | (1.28, 1.48) | (1.26, 1.38) | (1.38, 1.53) | (1.34, 1.49) | (1.18, 1.31) | (1.37, 1.55) | (1.41, 1.66) |
| Stratified by Race | | | | | | | | | | | |
| White | | | | | | | | | | | |
| n | 3,937 | 13,958 | 13,486 | 13,486 | 13,486 | 13,984 | 13,984 | 13,984 | 13,447 | 13,447 | 13,447 |
| BA | 1.44 | 1.54 | 1.27 | 1.47 | 1.34 | 1.39 | 1.44 | 1.47 | 1.28 | 1.49 | 1.52 |
| | (1.21, 1.72) | (1.47, 1.6) | (1.21, 1.33) | (1.39, 1.55) | (1.27, 1.42) | (1.33, 1.45) | (1.38, 1.51) | (1.4, 1.54) | (1.22, 1.34) | (1.42, 1.58) | (1.43, 1.63) |
| Black | | | | | | | | | | | |
| n | 1,467 | 5,176 | 4,887 | 4,887 | 4,887 | 5,201 | 5,201 | 5,201 | 4,851 | 4,851 | 4,851 |
| BA | 1.51 | 1.37 | 1.21 | 1.3 | 1.4 | 1.3 | 1.31 | 1.32 | 1.26 | 1.36 | 1.47 |
| | (1.15, 1.99) | (1.28, 1.47) | (1.12, 1.31) | (1.19, 1.42) | (1.27, 1.55) | (1.22, 1.38) | (1.22, 1.39) | (1.21, 1.44) | (1.17, 1.35) | (1.26, 1.48) | (1.31, 1.64) |
| Other | | | | | | | | | | | |
| n | 2,830 | 8,703 | 8,315 | 8,315 | 8,315 | 8,719 | 8,719 | 8,719 | 8,282 | 8,282 | 8,282 |
| BA | 1.21 | 1.37 | 1.18 | 1.25 | 1.22 | 1.27 | 1.32 | 1.27 | 1.19 | 1.3 | 1.27 |
| | (0.89, 1.65) | (1.28, 1.48) | (1.09, 1.28) | (1.14, 1.36) | (1.12, 1.34) | (1.19, 1.36) | (1.22, 1.42) | (1.18, 1.37) | (1.1, 1.29) | (1.19, 1.42) | (1.15, 1.4) |
| People Aged 65 and Younger | | | | | | | | | | | |
| ≤65 years | | | | | | | | | | | |
| n | 6,915 | 21,252 | 20,430 | 20,430 | 20,430 | 21,292 | 21,292 | 21,292 | 20,344 | 20,344 | 20,344 |
| BA | 1.27 | 1.61 | 1.29 | 1.47 | 1.45 | 1.46 | 1.51 | 1.49 | 1.34 | 1.51 | 1.54 |
| | (1.06, 1.53) | (1.52, 1.7) | (1.21, 1.39) | (1.38, 1.58) | (1.34, 1.56) | (1.39, 1.54) | (1.43, 1.6) | (1.4, 1.58) | (1.25, 1.44) | (1.41, 1.61) | (1.42, 1.67) |

Biological Age (BA) coefficients in the table are hazard ratios estimated from Cox proportional hazard regressions. KDM and PhenoAge measures were differenced from chronological age for analysis. These differenced values were then standardized to have M=0, SD=1 separately for men and women within the analysis sample so that effect-sizes are denominated in terms of a sex-specific 1 SD unit increase in BA advancement. The original KDM algorithm (left-most column) was projected onto data from NHANES 2007-2010 only because other NHANES IV waves did not include spirometry measurements. The original PhenoAge algorithm (second column from left) was projected onto data from NHANES 1999-2010 and 2015-2018 only because the intervening waves did not include CRP measurements

**Table S1.2** Associations of biological aging measures with healthspan-related characteristics in the NHANES IV

| | Original | | CALERIE Original | | | V1 | | | V2 | | |
|---|---|---|---|---|---|---|---|---|---|---|---|
| | KDM | PhenoAge | KDM | PhenoAge | HD (log) | KDM | PhenoAge | HD (log) | KDM | PhenoAge | HD (log) |
| | b (95% CI) | | | | | | | | | | |
| Full Sample | | | | | | | | | | | |
| SRH | 0.25 (0.23, 0.27) | 0.21 (0.2, 0.22) | 0.16 (0.15, 0.17) | 0.2 (0.19, 0.21) | 0.15 (0.13, 0.16) | 0.17 (0.16, 0.18) | 0.16 (0.15, 0.17) | 0.2 (0.19, 0.21) | 0.15 (0.14, 0.16) | 0.18 (0.17, 0.19) | 0.23 (0.22, 0.24) |
| ADL | 0.13 (0.1, 0.16) | 0.17 (0.15, 0.19) | 0.09 (0.07, 0.1) | 0.14 (0.12, 0.15) | 0.09 (0.08, 0.11) | 0.14 (0.12, 0.15) | 0.16 (0.14, 0.18) | 0.13 (0.12, 0.15) | 0.09 (0.08, 0.11) | 0.14 (0.13, 0.16) | 0.12 (0.11, 0.14) |
| Walk speed | - | 0.21 (0.18, 0.24) | 0.1 (0.08, 0.13) | 0.16 (0.14, 0.19) | 0.12 (0.09, 0.15) | 0.16 (0.13, 0.18) | 0.17 (0.14, 0.2) | 0.18 (0.15, 0.2) | 0.1 (0.07, 0.12) | 0.15 (0.12, 0.18) | 0.17 (0.14, 0.2) |
| Grip strength | - | - | - | - | - | - | - | - | - | - | - |
| Stratified by Gender | | | | | | | | | | | |
| Men | | | | | | | | | | | |
| SRH | 0.23 (0.2, 0.26) | 0.22 (0.2, 0.23) | 0.17 (0.15, 0.19) | 0.19 (0.17, 0.21) | 0.16 (0.14, 0.18) | 0.18 (0.16, 0.19) | 0.17 (0.15, 0.19) | 0.19 (0.17, 0.2) | 0.17 (0.15, 0.18) | 0.18 (0.17, 0.2) | 0.22 (0.2, 0.24) |
| ADL | 0.12 (0.08, 0.15) | 0.15 (0.13, 0.17) | 0.09 (0.07, 0.11) | 0.12 (0.1, 0.14) | 0.08 (0.06, 0.1) | 0.15 (0.13, 0.17) | 0.15 (0.13, 0.17) | 0.12 (0.1, 0.14) | 0.1 (0.08, 0.12) | 0.12 (0.1, 0.14) | 0.1 (0.08, 0.13) |
| Walk speed | - | 0.16 (0.12, 0.2) | 0.09 (0.05, 0.12) | 0.13 (0.09, 0.16) | 0.11 (0.07, 0.14) | 0.12 (0.09, 0.16) | 0.11 (0.08, 0.15) | 0.14 (0.11, 0.18) | 0.08 (0.05, 0.12) | 0.11 (0.07, 0.14) | 0.14 (0.1, 0.18) |
| Grip strength | - | - | - | - | - | - | - | - | - | - | - |
| Women | | | | | | | | | | | |
| SRH | 0.28 (0.25, 0.31) | 0.22 (0.2, 0.23) | 0.16 (0.14, 0.17) | 0.21 (0.2, 0.23) | 0.13 (0.11, 0.15) | 0.16 (0.14, 0.17) | 0.16 (0.14, 0.17) | 0.21 (0.2, 0.23) | 0.14 (0.12, 0.15) | 0.17 (0.15, 0.19) | 0.24 (0.23, 0.26) |
| ADL | 0.14 (0.09, 0.19) | 0.19 (0.17, 0.22) | 0.08 (0.06, 0.1) | 0.16 (0.13, 0.18) | 0.1 (0.08, 0.13) | 0.13 (0.11, 0.15) | 0.18 (0.15, 0.2) | 0.15 (0.13, 0.17) | 0.08 (0.06, 0.11) | 0.16 (0.14, 0.19) | 0.16 (0.13, 0.19) |
| Walk speed | - | 0.27 (0.22, 0.31) | 0.12 (0.08, 0.16) | 0.2 (0.16, 0.25) | 0.13 (0.08, 0.18) | 0.19 (0.15, 0.23) | 0.23 (0.18, 0.27) | 0.21 (0.17, 0.25) | 0.11 (0.07, 0.15) | 0.19 (0.14, 0.23) | 0.21 (0.16, 0.26) |
| Grip strength | - | - | - | - | - | - | - | - | - | - | - |
| Stratified by Race | | | | | | | | | | | |
| White | | | | | | | | | | | |
| SRH | 0.3 (0.27, 0.33) | 0.27 (0.25, 0.28) | 0.17 (0.15, 0.19) | 0.26 (0.25, 0.28) | 0.15 (0.14, 0.17) | 0.21 (0.2, 0.23) | 0.22 (0.21, 0.24) | 0.21 (0.2, 0.23) | 0.17 (0.15, 0.18) | 0.25 (0.23, 0.26) | 0.24 (0.23, 0.26) |
| ADL | 0.15 (0.11, 0.19) | 0.2 (0.18, 0.22) | 0.09 (0.07, 0.11) | 0.16 (0.14, 0.19) | 0.1 (0.08, 0.12) | 0.15 (0.13, 0.17) | 0.18 (0.16, 0.2) | 0.15 (0.13, 0.17) | 0.1 (0.08, 0.12) | 0.17 (0.15, 0.19) | 0.13 (0.11, 0.16) |
| Walk speed | - | 0.25 (0.21, 0.28) | 0.12 (0.08, 0.15) | 0.18 (0.15, 0.22) | 0.12 (0.08, 0.16) | 0.17 (0.13, 0.21) | 0.2 (0.17, 0.24) | 0.17 (0.13, 0.2) | 0.12 (0.09, 0.15) | 0.18 (0.15, 0.22) | 0.16 (0.12, 0.2) |
| Grip strength | - | - | - | - | - | - | - | - | - | - | - |
| Black | | | | | | | | | | | |

| | Original | | CALERIE Original | | | V1 | | | V2 | | |
|---|---|---|---|---|---|---|---|---|---|---|---|
| | KDM | PhenoAge | KDM | PhenoAge | HD (log) | KDM | PhenoAge | HD (log) | KDM | PhenoAge | HD (log) |
| | b (95% CI) | | | | | | | | | | |
| SRH | 0.18 (0.13, 0.22) | 0.17 (0.14, 0.19) | 0.15 (0.13, 0.17) | 0.17 (0.15, 0.2) | 0.15 (0.13, 0.18) | 0.14 (0.11, 0.16) | 0.14 (0.12, 0.16) | 0.17 (0.14, 0.19) | 0.15 (0.13, 0.18) | 0.16 (0.14, 0.18) | 0.21 (0.18, 0.24) |
| ADL | 0.08 (0, 0.16) | 0.13 (0.1, 0.17) | 0.07 (0.03, 0.11) | 0.1 (0.06, 0.14) | 0.1 (0.06, 0.14) | 0.13 (0.1, 0.17) | 0.14 (0.11, 0.17) | 0.14 (0.1, 0.18) | 0.08 (0.05, 0.12) | 0.11 (0.08, 0.15) | 0.12 (0.07, 0.17) |
| Walk speed | - | 0.14 (0.07, 0.21) | 0.02 (-0.05, 0.08) | 0.08 (0.01, 0.16) | 0.07 (-0.01, 0.15) | 0.11 (0.05, 0.18) | 0.13 (0.06, 0.19) | 0.15 (0.07, 0.22) | 0.03 (-0.03, 0.1) | 0.1 (0.03, 0.17) | 0.11 (0.02, 0.2) |
| Grip strength | - | - | - | - | - | - | - | - | - | - | - |
| Other | | | | | | | | | | | |
| SRH | 0.15 (0.11, 0.19) | 0.15 (0.13, 0.17) | 0.12 (0.1, 0.14) | 0.14 (0.13, 0.16) | 0.11 (0.09, 0.13) | 0.14 (0.12, 0.16) | 0.14 (0.12, 0.15) | 0.15 (0.13, 0.17) | 0.11 (0.09, 0.13) | 0.13 (0.11, 0.15) | 0.17 (0.15, 0.19) |
| ADL | 0.15 (0.09, 0.21) | 0.15 (0.12, 0.18) | 0.09 (0.06, 0.12) | 0.14 (0.11, 0.17) | 0.08 (0.05, 0.11) | 0.13 (0.1, 0.16) | 0.14 (0.11, 0.18) | 0.11 (0.08, 0.14) | 0.09 (0.06, 0.12) | 0.12 (0.09, 0.15) | 0.12 (0.08, 0.15) |
| Walk speed | - | 0.15 (0.09, 0.21) | 0.02 (-0.04, 0.07) | 0.09 (0.04, 0.15) | 0.04 (-0.01, 0.1) | 0.11 (0.06, 0.16) | 0.14 (0.09, 0.2) | 0.11 (0.06, 0.17) | 0.01 (-0.04, 0.07) | 0.08 (0.03, 0.14) | 0.07 (0.01, 0.14) |
| Grip strength | - | - | - | - | - | - | - | - | - | - | - |
| Stratified by Age | | | | | | | | | | | |
| Age 20-40 | | | | | | | | | | | |
| SRH | 0.21 (0.17, 0.25) | 0.19 (0.17, 0.21) | 0.15 (0.13, 0.17) | 0.16 (0.14, 0.18) | 0.07 (0.05, 0.09) | 0.11 (0.09, 0.14) | 0.09 (0.07, 0.12) | 0.12 (0.09, 0.14) | 0.15 (0.12, 0.17) | 0.12 (0.1, 0.14) | 0.15 (0.13, 0.18) |
| ADL | 0.02 (-0.08, 0.13) | 0.11 (0.06, 0.17) | 0.03 (-0.03, 0.08) | 0.07 (0.02, 0.13) | 0.03 (-0.02, 0.08) | 0.07 (0.02, 0.13) | 0.08 (0.03, 0.13) | 0.08 (0.02, 0.13) | 0.04 (-0.02, 0.1) | 0.07 (0.02, 0.12) | 0.06 (0, 0.12) |
| Walk speed | - | - | - | - | - | - | - | - | - | - | - |
| Grip strength | - | - | - | - | - | - | - | - | - | - | - |
| Age 40-60 | | | | | | | | | | | |
| SRH | 0.28 (0.25, 0.32) | 0.26 (0.24, 0.28) | 0.2 (0.18, 0.22) | 0.24 (0.22, 0.26) | 0.21 (0.19, 0.23) | 0.22 (0.2, 0.24) | 0.23 (0.21, 0.25) | 0.24 (0.22, 0.26) | 0.19 (0.17, 0.21) | 0.22 (0.2, 0.24) | 0.28 (0.26, 0.3) |
| ADL | 0.18 (0.09, 0.27) | 0.17 (0.12, 0.21) | 0.09 (0.05, 0.14) | 0.14 (0.1, 0.19) | 0.14 (0.09, 0.19) | 0.13 (0.08, 0.18) | 0.14 (0.1, 0.19) | 0.15 (0.11, 0.2) | 0.09 (0.05, 0.14) | 0.12 (0.08, 0.17) | 0.17 (0.12, 0.22) |
| Walk speed | - | 0.22 (0.17, 0.27) | 0.15 (0.11, 0.2) | 0.19 (0.14, 0.23) | 0.18 (0.14, 0.23) | 0.17 (0.13, 0.22) | 0.18 (0.14, 0.23) | 0.2 (0.16, 0.24) | 0.15 (0.1, 0.19) | 0.17 (0.13, 0.22) | 0.22 (0.17, 0.26) |
| Grip strength | - | - | - | - | - | - | - | - | - | - | - |
| Age 60-80 | | | | | | | | | | | |
| SRH | 0.26 (0.22, 0.3) | 0.21 (0.19, 0.23) | 0.15 (0.13, 0.16) | 0.21 (0.19, 0.23) | 0.17 (0.15, 0.19) | 0.17 (0.15, 0.18) | 0.18 (0.16, 0.2) | 0.22 (0.2, 0.24) | 0.14 (0.12, 0.15) | 0.2 (0.18, 0.22) | 0.26 (0.24, 0.28) |
| ADL | 0.11 (0.08, 0.15) | 0.16 (0.14, 0.17) | 0.09 (0.07, 0.11) | 0.14 (0.12, 0.15) | 0.09 (0.07, 0.1) | 0.13 (0.12, 0.15) | 0.15 (0.13, 0.16) | 0.13 (0.11, 0.14) | 0.09 (0.07, 0.11) | 0.14 (0.12, 0.16) | 0.12 (0.1, 0.15) |
| Walk speed | - | 0.21 | 0.1 | 0.16 | 0.11 | 0.15 | 0.16 | 0.16 | 0.09 | 0.14 | 0.16 |

| | Original | | CALERIE Original | | | V1 | | | V2 | | |
|---|---|---|---|---|---|---|---|---|---|---|---|
| | KDM | PhenoAge | KDM | PhenoAge | HD (log) | KDM | PhenoAge | HD (log) | KDM | PhenoAge | HD (log) |
| | | | | | | b (95% CI) | | | | | |
| | | (0.17, 0.25) | (0.06, 0.13) | (0.12, 0.19) | (0.07, 0.15) | (0.12, 0.19) | (0.12, 0.2) | (0.13, 0.2) | (0.06, 0.13) | (0.1, 0.18) | (0.12, 0.2) |
| Grip strength | - | - | - | - | - | - | - | - | - | - | - |

Coefficients are from linear regressions of healthspan-related characteristics on biological aging measures. Outcome variables were standardized to have M=0, SD=1 for analysis. Standardization was performed separately for men and women in the case of grip strength. Walk speed was log transformed prior to standardization to reduce skew. KDM and PhenoAge measures were differenced from chronological age for analysis (i.e. values = BA-CA). These differenced values were then standardized to have M=0, SD=1 separately for men and women within the analysis sample so that effect-sizes are denominated in terms of a sex-specific 1 SD unit increase in biological age advancement. Models included covariates for chronological age and sex. The original KDM algorithm (left-most column) was projected onto data from NHANES 2007-2010 only because other NHANES IV waves did not include spirometry measurements. The original PhenoAge algorithm (second column from left) was projected onto data from NHANES 1999-2010 and 2015-2018 only because the intervening waves did not include CRP measurements. Walk speed was measured only in NHANES 1999-2002 and is available only for participants aged 50 and older. Grip strength was measured only in NHANES 2011-2014

**Table S1.3** Associations of socioeconomic circumstances with biological aging measures in the NHANES IV

| | Original | | CALERIE Original | | | V1 | | | V2 | | |
|---|---|---|---|---|---|---|---|---|---|---|---|
| | KDM | PhenoAge | KDM | PhenoAge | HD (log) | KDM | PhenoAge | HD (log) | KDM | PhenoAge | HD (log) |
| | b (95% CI) | | | | | | | | | | |
| Full Sample | | | | | | | | | | | |
| Education | -0.19<br>(-0.22, -0.17) | -0.07<br>(-0.08, -0.06) | -0.08<br>(-0.09, -0.07) | -0.1<br>(-0.11, -0.09) | -0.1<br>(-0.11, -0.09) | -0.07<br>(-0.08, -0.06) | -0.07<br>(-0.08, -0.06) | -0.09<br>(-0.1, -0.08) | -0.07<br>(-0.08, -0.06) | -0.09<br>(-0.1, -0.07) | -0.11<br>(-0.12, -0.1) |
| Annual income | -0.17<br>(-0.19, -0.15) | -0.11<br>(-0.12, -0.1) | -0.07<br>(-0.08, -0.06) | -0.13<br>(-0.14, -0.12) | -0.12<br>(-0.13, -0.11) | -0.1<br>(-0.11, -0.09) | -0.1<br>(-0.11, -0.09) | -0.1<br>(-0.11, -0.09) | -0.08<br>(-0.09, -0.07) | -0.12<br>(-0.13, -0.11) | -0.11<br>(-0.12, -0.11) |
| Poverty ratio | -0.18<br>(-0.21, -0.16) | -0.14<br>(-0.15, -0.13) | -0.07<br>(-0.08, -0.06) | -0.13<br>(-0.14, -0.12) | -0.1<br>(-0.11, -0.09) | -0.09<br>(-0.1, -0.08) | -0.09<br>(-0.1, -0.08) | -0.11<br>(-0.12, -0.1) | -0.07<br>(-0.08, -0.06) | -0.11<br>(-0.12, -0.1) | -0.12<br>(-0.13, -0.11) |
| Stratified by Gender | | | | | | | | | | | |
| Men | | | | | | | | | | | |
| Education | -0.21<br>(-0.24, -0.18) | -0.08<br>(-0.09, -0.06) | -0.08<br>(-0.1, -0.07) | -0.1<br>(-0.11, -0.08) | -0.09<br>(-0.1, -0.07) | -0.07<br>(-0.09, -0.06) | -0.08<br>(-0.09, -0.06) | -0.07<br>(-0.09, -0.06) | -0.08<br>(-0.09, -0.06) | -0.09<br>(-0.11, -0.08) | -0.1<br>(-0.11, -0.09) |
| Annual income | -0.16<br>(-0.19, -0.13) | -0.1<br>(-0.11, -0.08) | -0.05<br>(-0.07, -0.03) | -0.11<br>(-0.12, -0.09) | -0.11<br>(-0.13, -0.1) | -0.09<br>(-0.11, -0.07) | -0.09<br>(-0.11, -0.08) | -0.09<br>(-0.1, -0.07) | -0.06<br>(-0.08, -0.05) | -0.11<br>(-0.13, -0.1) | -0.1<br>(-0.12, -0.09) |
| Poverty ratio | -0.18<br>(-0.21, -0.15) | -0.12<br>(-0.13, -0.1) | -0.07<br>(-0.08, -0.05) | -0.12<br>(-0.13, -0.1) | -0.1<br>(-0.12, -0.09) | -0.09<br>(-0.1, -0.07) | -0.09<br>(-0.1, -0.07) | -0.1<br>(-0.11, -0.08) | -0.07<br>(-0.08, -0.05) | -0.11<br>(-0.12, -0.09) | -0.11<br>(-0.12, -0.1) |
| Women | | | | | | | | | | | |
| Education | -0.18<br>(-0.21, -0.15) | -0.08<br>(-0.1, -0.07) | -0.07<br>(-0.09, -0.06) | -0.11<br>(-0.12, -0.09) | -0.1<br>(-0.12, -0.09) | -0.07<br>(-0.08, -0.05) | -0.08<br>(-0.09, -0.06) | -0.12<br>(-0.13, -0.1) | -0.06<br>(-0.08, -0.05) | -0.08<br>(-0.1, -0.07) | -0.12<br>(-0.13, -0.1) |
| Annual income | -0.18<br>(-0.21, -0.15) | -0.12<br>(-0.13, -0.11) | -0.08<br>(-0.1, -0.07) | -0.15<br>(-0.17, -0.14) | -0.12<br>(-0.14, -0.11) | -0.11<br>(-0.12, -0.09) | -0.11<br>(-0.13, -0.1) | -0.12<br>(-0.13, -0.11) | -0.09<br>(-0.1, -0.07) | -0.13<br>(-0.15, -0.12) | -0.12<br>(-0.14, -0.11) |
| Poverty ratio | -0.19<br>(-0.22, -0.16) | -0.15<br>(-0.17, -0.14) | -0.08<br>(-0.09, -0.06) | -0.15<br>(-0.16, -0.13) | -0.1<br>(-0.11, -0.08) | -0.09<br>(-0.1, -0.08) | -0.09<br>(-0.11, -0.08) | -0.13<br>(-0.15, -0.12) | -0.07<br>(-0.08, -0.05) | -0.11<br>(-0.13, -0.1) | -0.12<br>(-0.13, -0.11) |
| Stratified by Race | | | | | | | | | | | |
| White | | | | | | | | | | | |
| Education | -0.24<br>(-0.27, -0.21) | -0.14<br>(-0.15, -0.12) | -0.1<br>(-0.11, -0.08) | -0.19<br>(-0.21, -0.18) | -0.1<br>(-0.11, -0.08) | -0.12<br>(-0.13, -0.1) | -0.15<br>(-0.16, -0.13) | -0.1<br>(-0.12, -0.09) | -0.1<br>(-0.11, -0.08) | -0.17<br>(-0.19, -0.16) | -0.12<br>(-0.13, -0.1) |
| Annual income | -0.18<br>(-0.21, -0.15) | -0.14<br>(-0.16, -0.13) | -0.07<br>(-0.09, -0.06) | -0.18<br>(-0.19, -0.16) | -0.12<br>(-0.13, -0.1) | -0.11<br>(-0.13, -0.1) | -0.14<br>(-0.16, -0.13) | -0.11<br>(-0.12, -0.09) | -0.09<br>(-0.1, -0.07) | -0.17<br>(-0.18, -0.15) | -0.12<br>(-0.13, -0.11) |
| Poverty ratio | -0.19<br>(-0.21, -0.16) | -0.17<br>(-0.19, -0.16) | -0.08<br>(-0.09, -0.06) | -0.19<br>(-0.2, -0.17) | -0.1<br>(-0.11, -0.08) | -0.12<br>(-0.13, -0.1) | -0.14<br>(-0.16, -0.13) | -0.11<br>(-0.13, -0.1) | -0.08<br>(-0.1, -0.07) | -0.17<br>(-0.18, -0.15) | -0.11<br>(-0.13, -0.1) |
| Black | | | | | | | | | | | |
| Education | -0.11<br>(-0.17, -0.06) | -0.04<br>(-0.06, -0.01) | -0.03<br>(-0.05, 0) | -0.06<br>(-0.08, -0.03) | -0.09<br>(-0.11, -0.07) | -0.05<br>(-0.08, -0.03) | -0.06<br>(-0.09, -0.04) | -0.05<br>(-0.07, -0.02) | -0.05<br>(-0.08, -0.02) | -0.08<br>(-0.11, -0.05) | -0.06<br>(-0.08, -0.04) |
| Annual income | -0.12<br>(-0.18, -0.07) | -0.1<br>(-0.12, -0.07) | -0.07<br>(-0.1, -0.05) | -0.08<br>(-0.11, -0.06) | -0.12<br>(-0.14, -0.1) | -0.11<br>(-0.13, -0.08) | -0.1<br>(-0.13, -0.08) | -0.07<br>(-0.1, -0.05) | -0.1<br>(-0.13, -0.08) | -0.11<br>(-0.14, -0.09) | -0.08<br>(-0.1, -0.06) |
| Poverty ratio | -0.12<br>(-0.17, -0.06) | -0.14<br>(-0.16, -0.11) | -0.08<br>(-0.1, -0.05) | -0.1<br>(-0.12, -0.07) | -0.11<br>(-0.14, -0.09) | -0.09<br>(-0.12, -0.07) | -0.09<br>(-0.12, -0.07) | -0.08<br>(-0.1, -0.06) | -0.1<br>(-0.12, -0.07) | -0.11<br>(-0.14, -0.09) | -0.09<br>(-0.11, -0.07) |
| Other | | | | | | | | | | | |
| Education | -0.1<br>(-0.13, -0.07) | -0.02<br>(-0.03, 0) | -0.04<br>(-0.06, -0.03) | -0.04<br>(-0.06, -0.03) | -0.09<br>(-0.11, -0.08) | -0.05<br>(-0.07, -0.04) | -0.05<br>(-0.07, -0.03) | -0.07<br>(-0.08, -0.05) | -0.04<br>(-0.06, -0.03) | -0.05<br>(-0.07, -0.03) | -0.08<br>(-0.1, -0.07) |

| | Original | | CALERIE Original | | | V1 | | | V2 | | |
|---|---|---|---|---|---|---|---|---|---|---|---|
| | KDM | PhenoAge | KDM | PhenoAge | HD (log) | KDM | PhenoAge | HD (log) | KDM | PhenoAge | HD (log) |
| | | | | | | b (95% CI) | | | | | |
| Annual income | -0.09 (-0.13, -0.05) | -0.03 (-0.05, -0.02) | -0.02 (-0.04, -0.0002) | -0.07 (-0.09, -0.05) | -0.11 (-0.12, -0.09) | -0.07 (-0.09, -0.05) | -0.08 (-0.1, -0.06) | -0.08 (-0.09, -0.06) | -0.03 (-0.05, -0.02) | -0.09 (-0.1, -0.07) | -0.08 (-0.1, -0.06) |
| Poverty ratio | -0.11 (-0.15, -0.08) | -0.06 (-0.08, -0.05) | -0.02 (-0.04, -0.003) | -0.07 (-0.09, -0.05) | -0.09 (-0.11, -0.07) | -0.06 (-0.08, -0.04) | -0.07 (-0.09, -0.05) | -0.09 (-0.11, -0.07) | -0.02 (-0.04, -0.01) | -0.07 (-0.09, -0.05) | -0.08 (-0.1, -0.06) |
| Stratified by Age | | | | | | | | | | | |
| Age 20-40 | | | | | | | | | | | |
| Education | -0.14 (-0.18, -0.11) | -0.08 (-0.09, -0.06) | -0.03 (-0.04, -0.01) | -0.07 (-0.09, -0.06) | -0.04 (-0.05, -0.02) | -0.05 (-0.06, -0.03) | -0.07 (-0.08, -0.05) | -0.06 (-0.08, -0.05) | -0.02 (-0.04, -0.01) | -0.06 (-0.08, -0.04) | -0.07 (-0.09, -0.06) |
| Annual income | -0.09 (-0.12, -0.06) | -0.06 (-0.08, -0.05) | 0.001 (-0.01, 0.02) | -0.06 (-0.08, -0.04) | -0.03 (-0.04, -0.01) | -0.04 (-0.06, -0.03) | -0.04 (-0.06, -0.03) | -0.04 (-0.06, -0.03) | -0.004 (-0.02, 0.01) | -0.05 (-0.07, -0.03) | -0.05 (-0.06, -0.03) |
| Poverty ratio | -0.13 (-0.16, -0.1) | -0.1 (-0.11, -0.08) | -0.02 (-0.04, -0.005) | -0.07 (-0.09, -0.05) | -0.004 (-0.02, 0.01) | -0.04 (-0.05, -0.02) | -0.03 (-0.05, -0.02) | -0.05 (-0.07, -0.04) | -0.01 (-0.02, 0.01) | -0.04 (-0.05, -0.02) | -0.05 (-0.07, -0.04) |
| Age 40-60 | | | | | | | | | | | |
| Education | -0.2 (-0.24, -0.17) | -0.09 (-0.1, -0.07) | -0.1 (-0.11, -0.08) | -0.1 (-0.12, -0.08) | -0.11 (-0.13, -0.09) | -0.08 (-0.09, -0.06) | -0.08 (-0.09, -0.06) | -0.11 (-0.12, -0.09) | -0.08 (-0.1, -0.07) | -0.09 (-0.11, -0.07) | -0.13 (-0.14, -0.11) |
| Annual income | -0.19 (-0.23, -0.16) | -0.15 (-0.17, -0.13) | -0.09 (-0.11, -0.07) | -0.17 (-0.19, -0.15) | -0.14 (-0.15, -0.12) | -0.13 (-0.14, -0.11) | -0.13 (-0.15, -0.11) | -0.14 (-0.16, -0.12) | -0.09 (-0.11, -0.08) | -0.16 (-0.18, -0.14) | -0.15 (-0.17, -0.13) |
| Poverty ratio | -0.2 (-0.23, -0.16) | -0.17 (-0.18, -0.15) | -0.08 (-0.1, -0.07) | -0.15 (-0.17, -0.13) | -0.12 (-0.13, -0.1) | -0.11 (-0.12, -0.09) | -0.11 (-0.12, -0.09) | -0.14 (-0.16, -0.12) | -0.08 (-0.09, -0.06) | -0.13 (-0.15, -0.11) | -0.14 (-0.16, -0.13) |
| Age 60-80 | | | | | | | | | | | |
| Education | -0.24 (-0.28, -0.2) | -0.07 (-0.09, -0.05) | -0.1 (-0.13, -0.08) | -0.12 (-0.14, -0.1) | -0.12 (-0.14, -0.1) | -0.07 (-0.09, -0.05) | -0.06 (-0.08, -0.04) | -0.1 (-0.12, -0.08) | -0.09 (-0.11, -0.07) | -0.09 (-0.11, -0.07) | -0.12 (-0.14, -0.1) |
| Annual income | -0.24 (-0.29, -0.19) | -0.1 (-0.12, -0.08) | -0.12 (-0.14, -0.1) | -0.15 (-0.18, -0.13) | -0.16 (-0.18, -0.14) | -0.11 (-0.13, -0.09) | -0.11 (-0.13, -0.08) | -0.12 (-0.14, -0.1) | -0.11 (-0.14, -0.09) | -0.13 (-0.15, -0.11) | -0.14 (-0.16, -0.12) |
| Poverty ratio | -0.25 (-0.3, -0.21) | -0.14 (-0.16, -0.12) | -0.13 (-0.16, -0.11) | -0.17 (-0.19, -0.15) | -0.16 (-0.18, -0.14) | -0.12 (-0.14, -0.09) | -0.11 (-0.13, -0.08) | -0.14 (-0.17, -0.12) | -0.12 (-0.14, -0.09) | -0.14 (-0.16, -0.11) | -0.16 (-0.18, -0.14) |

Coefficients are from linear regressions of biological aging measures on measures of socioeconomic circumstances. KDM and PhenoAge measures were differenced from chronological age for analysis (i.e. values = BA-CA). These differenced values were then standardized to have M=0, SD=1 separately for men and women within the analysis sample. Socioeconomic circumstances measures were standardized to M=0, SD=1 for analysis so that effect-sizes are denominated in terms of a 1 SD unit improvement in socioeconomic circumstances. Models included covariates for chronological age and sex. The original KDM algorithm (left-most column) was projected onto data from NHANES 2007-2010 only because other NHANES IV waves did not include spirometry measurements. The original PhenoAge algorithm (second column from left) was projected onto data from NHANES 1999-2010 and 2015-2018 only because the intervening waves did not include CRP measurements

*II. Analysis of the CALERIE data using the BioAge package*

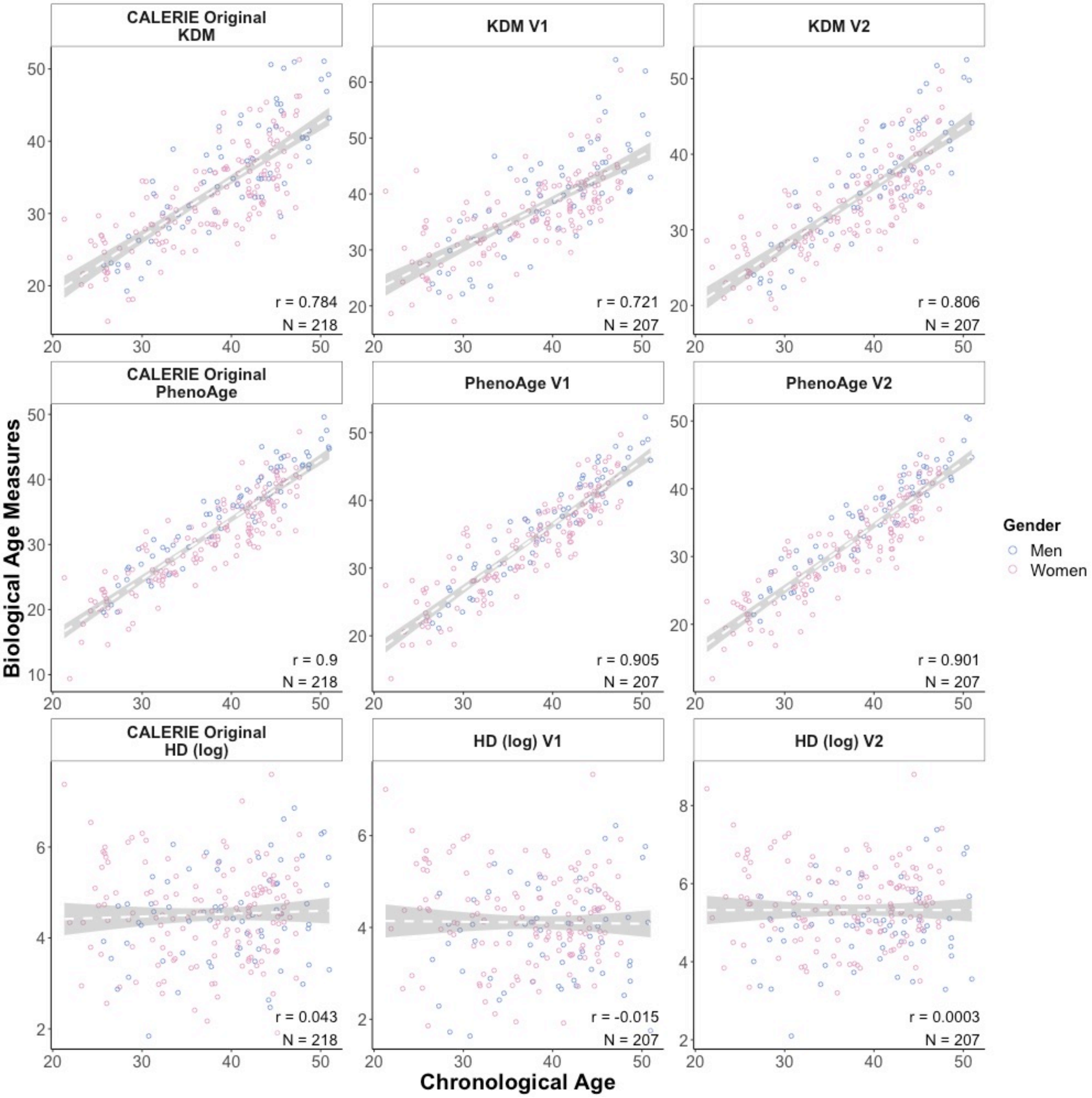


**Fig. S2.1** KDM, PhenoAge, and HD biological aging measures plotted against chronological age for participants in the CALERIE

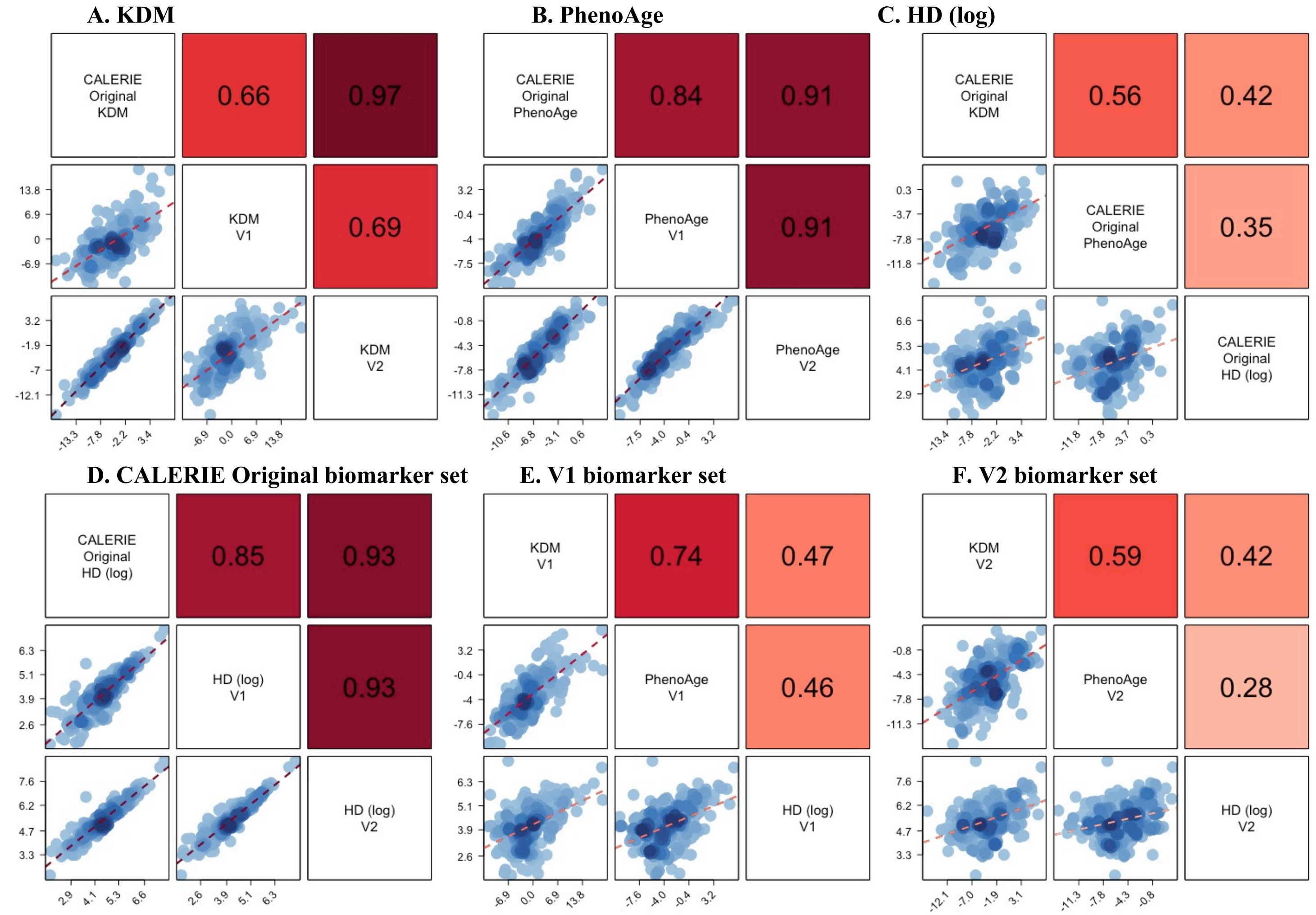


**Fig. S2.2** A matrix of association plots among KDM, PhenoAge, and HD biological aging measures in the CALERIE

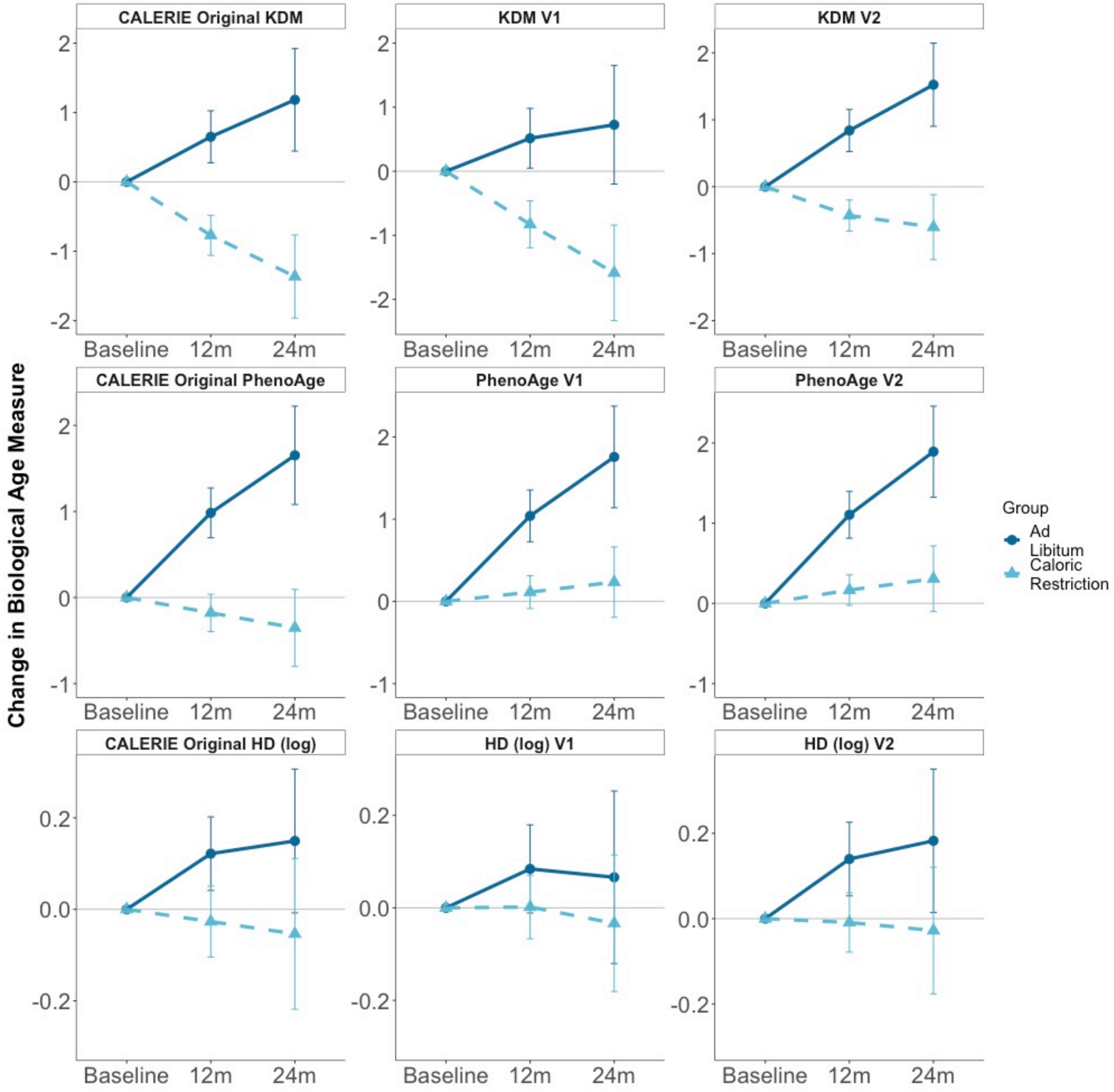


**Fig. S2.3** Change in KDM, PhenoAge, and HD biological aging measures from Baseline to 12- and 24-month follow-ups in the ad libitum (dark blue dots) and caloric-restriction (light blue triangles) arms of the CALERIE trial. Mean values with 95% confidence intervals were calculated for each follow-up

**Table S2.1** Parameters used to calculate biological aging measures in the CALRIE dataset

Panel A. Biomarker sets used to calculate the biological aging measures

| | Biomarker Set | | |
|---|---|---|---|
| | **CALERIE Original** | **V1** | **V2** |
| Biomarkers included in NHANES III Training Analysis | | | |
| Albumin | X | X | X |
| Alkaline Phosphatase | X | X | X |
| Blood Urea Nitrogen (BUN) | X | | X |
| Creatinine | X | X | X |
| C-reactive Protein (CRP) | X | X | X |
| Glucose | | X | |
| Glycated Hemoglobin (HbA1C) | X | | X |
| Lymphocyte % | | X | X |
| Mean Cell Volume (MCV) | | X | X |
| Red Cell Distribution Width (RDW) | | | |
| Systolic Blood Pressure (SBP) | X | | X |
| Total Cholesterol | X | | X |
| Uric Acid | X | | X |
| White Blood Cell Count (WBC) | X | X | X |
| Correlation with Chronological Age in NHANES IV | | | |
| KDM | 0.92 | 0.90 | 0.94 |
| PhenoAge | 0.97 | 0.97 | 0.97 |
| HD (log) | 0.35 | 0.28 | 0.50 |
| Correlation with Chronological Age in CALERIE | | | |
| KDM | 0.78 | 0.72 | 0.81 |
| PhenoAge | 0.90 | 0.91 | 0.90 |
| HD (log) | 0.04 | -0.02 | 0.00 |

Panel B. The NHANESIII (A) and CALERIE baseline (B) correlation matrices for the V2 biomarker sets. The correlation matrices include the set of biomarkers used to compose the V2 biological age algorithms, chronological age, the V2 biological age advance measures, and V2 homeostatic dysregulation. Biomarker abbreviations are as follows: ALP – alkaline phosphatase, CRP – C-reactive protein, HbA1C – glycated hemoglobin, SBP – systolic blood pressure, BUN – blood urea nitrogen, UAP – uric acid, MCV – mean cell volume, WBC – white blood cell count, CA – chronological age. Biological age advance values were computed by taking the difference of each of KDM Biological Age and PhenoAge from chronological age

**A. NHANESIII**

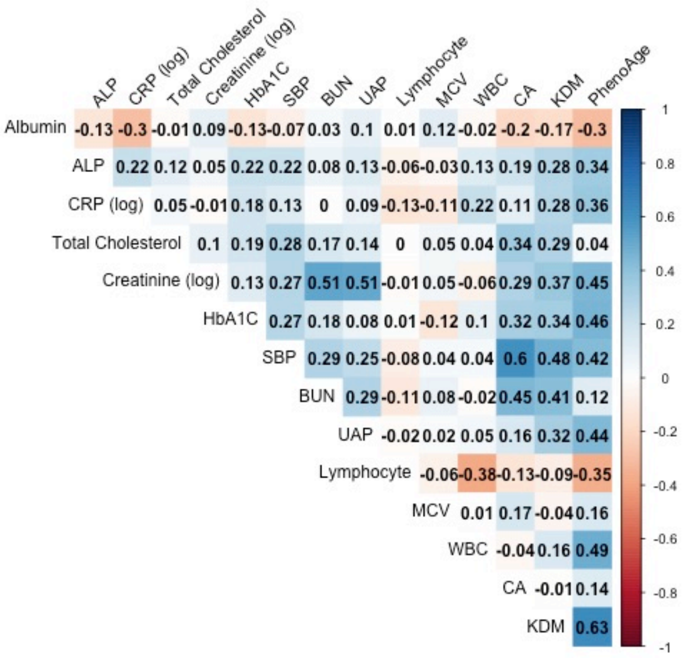


**B. CALERIE baseline**

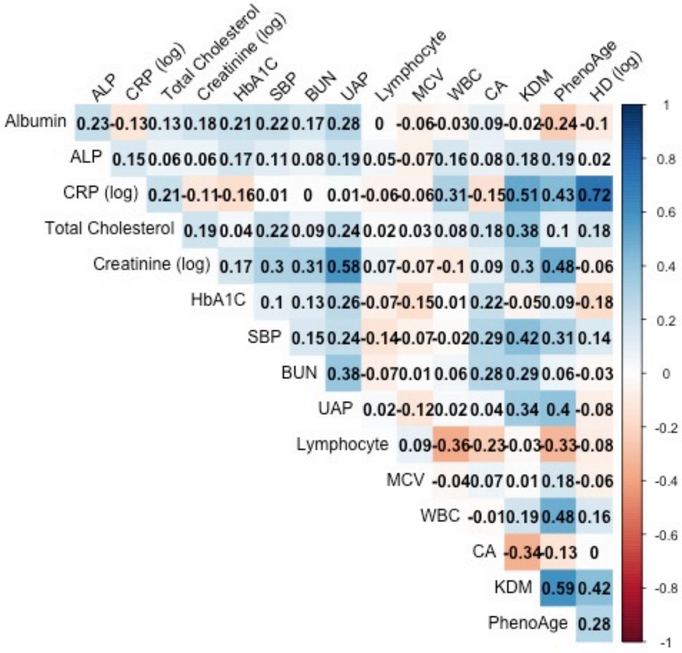

**Table S2.2** Summary statistics of biological aging measures in the CALERIE baseline

| Biomarker set | n | Mean | SD |
|---|---|---|---|
| Chronological age | 220 | 38.10 | 7.18 |
| CALERIE Original | | | |
| KDM | 218 | 33.25 | 7.28 |
| PhenoAge | 218 | 32.19 | 7.33 |
| HD (log) | 218 | 4.52 | 1.03 |
| V1 | | | |
| KDM | 207 | 37.17 | 7.95 |
| PhenoAge | 207 | 34.46 | 7.47 |
| HD (log) | 207 | 4.11 | 1.03 |
| V2 | | | |
| KDM | 207 | 34.17 | 6.96 |
| PhenoAge | 207 | 32.64 | 7.42 |
| HD (log) | 207 | 5.32 | 1.04 |

**Table S2.3** Estimated annual change in KDM, PhenoAge, and HD biological aging measures from baseline through 24-month follow-up in Ad libitum- and Caloric Restriction-arm Participants in the CALERIE Randomized Trial

| Biomarker Set | | b | [95% CI] | p value | n |
|---|---|---|---|---|---|
| CALERIE Original | | | | | |
| KDM | Ad libitum | 0.52 | [0, 1.05] | 0.053 | 73 |
| | Caloric restriction | -0.54 | [-0.92, -0.15] | 0.007 | 145 |
| | Interaction | -1.06 | [-1.71, -0.41] | 0.002 | 218 |
| PhenoAge | Ad libitum | 0.71 | [0.34, 1.09] | <0.01 | 73 |
| | Caloric restriction | -0.11 | [-0.39, 0.16] | 0.422 | 145 |
| | Interaction | -0.83 | [-1.29, -0.36] | 0.001 | 218 |
| HD (log) | Ad libitum | 0.05 | [-0.08, 0.18] | 0.447 | 73 |
| | Caloric restriction | -0.02 | [-0.12, 0.08] | 0.658 | 145 |
| | Interaction | -0.07 | [-0.24, 0.09] | 0.382 | 218 |
| V1 | | | | | |
| KDM | Ad libitum | 0.24 | [-0.44, 0.93] | 0.487 | 68 |
| | Caloric restriction | -0.66 | [-1.16, -0.16] | 0.01 | 139 |
| | Interaction | -0.9 | [-1.75, -0.06] | 0.037 | 207 |
| PhenoAge | Ad libitum | 0.77 | [0.38, 1.17] | <0.01 | 68 |
| | Caloric restriction | 0.13 | [-0.15, 0.42] | 0.372 | 139 |
| | Interaction | -0.64 | [-1.13, -0.16] | 0.01 | 207 |
| HD (log) | Ad libitum | 0 | [-0.14, 0.14] | 0.987 | 68 |
| | Caloric restriction | -0.02 | [-0.12, 0.08] | 0.72 | 139 |
| | Interaction | -0.02 | [-0.19, 0.15] | 0.823 | 207 |
| V2 | | | | | |
| KDM | Ad libitum | 0.69 | [0.21, 1.16] | 0.005 | 68 |
| | Caloric restriction | -0.2 | [-0.55, 0.14] | 0.248 | 139 |
| | Interaction | -0.89 | [-1.47, -0.31] | 0.003 | 207 |
| PhenoAge | Ad libitum | 0.83 | [0.46, 1.21] | <0.01 | 68 |
| | Caloric restriction | 0.17 | [-0.11, 0.44] | 0.232 | 139 |
| | Interaction | -0.67 | [-1.13, -0.2] | 0.006 | 207 |
| HD (log) | Ad libitum | 0.06 | [-0.08, 0.2] | 0.411 | 68 |
| | Caloric restriction | -0.01 | [-0.12, 0.09] | 0.784 | 139 |
| | Interaction | -0.07 | [-0.25, 0.1] | 0.409 | 207 |

The regression model included sex and age at baseline as covariates. CI = Confidence interval
